# Practice-Based Insights into Adult Genetic Medicine: High Diagnostic Yield, Indication-Specific Results, and Patterns of Test Utilization across 10,000 Patient Encounters

**DOI:** 10.1101/2025.10.09.25337579

**Authors:** Jessica I Gold, Yehuda Elkaim, Stephanie Asher, Zoe Bogus, Teresa Chai, Stacey Cohen, Courtney Condit, Isaac Elysee, Brielle N. Gehringer, Shannon M Gray, Laura Hennessy, Emma Kennedy, Anna Raper, Maria Bonanni, Ala Streater, Eamon Toye, Colleen Kripke, Katherine L Nathanson, Mersedeh Rohanizadegan, Staci Kallish, Theodore G Drivas

**Affiliations:** Division of Clinical Genetics, Department of Pediatrics, Cohen Children’s Medical Center, Northwell Health, Great Neck, NY, 11021, USA; Institute of Health System Science, Feinstein Institutes for Medical Research, Northwell Health, Manhasset, NY, 11030, USA; Perelman School of Medicine at the University of Pennsylvania, Philadelphia, PA, 19104, USA; Division of Translational Medicine and Human Genetics, Department of Medicine, Perelman School of Medicine at the University of Pennsylvania, Philadelphia, PA, 19104, USA; Kaiser Permanente, Tysons Corners Medical Center, McLean, VA, 22102, USA; Abramson Cancer Center, Perelman School of Medicine at the University of Pennsylvania, Philadelphia, PA, 19104, USA; Department of Genetics, Perelman School of Medicine at the University of Pennsylvania, Philadelphia, PA, 19104, USA

## Abstract

Genetic medicine has become a routine component of clinical care, yet the evidence guiding its implementation remains heavily weighted toward pediatric populations, with limited practice-level data to inform adult referral pathways, test selection, or service delivery. We analyzed eight years of electronic health record (EHR)-linked data from a high-volume adult genetics clinic (9,867 visits) within a major academic health system to define referral patterns, test utilization, and genetic testing outcomes across indications and demographic groups. While most visits were in-person and led by a genetics-trained physician, genetic counselor-only visits accounted for 12% of all encounters and telemedicine remained a substantial component of follow-up care, comprising 39% of follow-up visits from 2023 onward. Genetic testing was ordered for 52% of all new patients, with significant indication-specific variation in ordering propensity. Overall, 24% of all tests returned a diagnostic result. Diagnostic yield differed markedly by testing modality: exome sequencing yielded diagnoses in 40% of patients, whereas next-generation sequencing panels, the most frequently ordered testing type, yielded diagnoses in only 16%. Although diagnostic yield declined with increasing patient age, it remained above 17% in every age stratum, with the yield of exome/genome sequencing remaining above 30% across all age groups. Outcomes varied significantly by referral indication, with some indications yielding significantly high rates of pathogenic findings, and others more often yielding negative or uncertain results. Operational factors significantly shaped practice: test type and laboratory utilization shifted significantly over time in step with payer and testing lab policy changes. Together, these practice-based data establish adult genetics as a high-yield and important clinical domain. Our findings provide actionable evidence to inform age- and indication-aware triage, guide workforce planning, promote adult-focused practice guidelines, better inform payor reimbursement policies, and advance the integration of genomic medicine into routine adult patient care.

## Introduction

As genetic testing has become easier to order, broader in scope, and more clinically useful, it has moved from a specialized service to become a routine part of clinical medicine. In a recent electronic health record study of 1.8 million patients at a major academic health center, the proportion of patients with documented genetic testing rose six-fold between 2002 and 2022, multigene panels became the dominant testing modality, and the number of distinct diseases identified by genetic testing increased ten-fold.^1^ Genetic information now informs diagnosis, risk assessment, surveillance, treatment selection, pharmacogenomics, reproductive counseling, and cascade testing across organ systems and medical specialties.^2^

However, studies examining the use of genetic testing for the diagnostic evaluation of suspected genetic disorders have focused predominantly on pediatric cohorts with neurodevelopmental disorders and congenital anomalies. Adults remain underrepresented in this literature, limiting the evidence available to guide testing decisions in routine adult genetics care.^3,4^ Professional guidance reflects this history – pediatric genetics has accumulated recommendations covering several major parts of care: a joint American Academy of Pediatrics (AAP)/American College of Genetics and Genomics (ACMG) policy framework for diagnostic and predictive genetic testing in children;^5^ an established newborn-screening system; first-tier testing recommendations for exome or genome sequencing in developmental delay, intellectual disability, autism, and congenital anomalies;^6–8^ and numerous disease-specific pediatric health-supervision guidelines, such as the joint AAP/ACMG report for Neurofibromatosis type 1 (NF1) and AAP guidance for Marfan syndrome.^9,10^ These recommendations do not constitute a single end-to-end framework, but they provide substantially more structure for the implementation of genetic medicine in pediatric populations than is available in adult general genetics.

Adult guidance is not absent, but its scope varies substantially across clinical indications. Cancer genetics is a notable area of well-developed guidance, with NCCN and ASCO recommendations addressing eligibility for germline testing, test selection, and management of hereditary cancer risk. ACMG has also published general genetics referral indications for both pediatric and adult patients,^11^ and disease- or subspecialty-specific recommendations exist for inherited cardiac disease, familial hypercholesterolemia, and selected syndromes.^12,13^ Broader guidance for adult practice is less comprehensive: a major statement directed toward internists emphasizes the ethical and professional issues surrounding genetic testing rather than providing detailed recommendations for referral and test selection.^2^ As a result, despite established pathways for cancer and selected other conditions, evidence remains limited to guide referral prioritization, test selection, and counseling about expected diagnostic yield across the diverse presentations encountered in adult general genetics practice.^14,15^

Studies of diagnostic genetic testing in adults demonstrate substantial clinical value across a broad range of presentations. Genetic disease in adults can be difficult to recognize: penetrance is variable, family histories are often incomplete, and clinical features may resemble common acquired conditions. Nevertheless, studies of adults evaluated in genetics clinics for suspected genetic disease have reported diagnostic yields of 17%–29% with exome or genome sequencing.^4,16–18^ These diagnoses were clinically meaningful, frequently changing management and prompting cascade family testing.^16^ Genetic testing also is informative in routine adult specialty care in patients without obvious clinical suspicion of genetic disease. In a large cohort of patients with chronic kidney disease, 91.6% of whom were adults, exome sequencing identified a genetic diagnosis in 9.3% of all patients.^19^ Similarly, exome sequencing in critically ill adults aged 18-40 years identified a diagnostic genetic finding in 24% of patients, often with implications for care.^20^ Together, these studies demonstrate the diagnostic and clinical value of genetic testing across adult medicine. However, longitudinal data linking patient demographics and referral indications to testing decisions, diagnostic outcomes, and models of care delivery remain limited across the diverse population evaluated in adult general genetics clinics.

Published longitudinal descriptions of adult general genetics practice remain limited. Two major clinic series evaluated 1,552 adult patients over seven years and 30,780 visits across 39 years of available data, respectively.^21,22^ These studies characterized referral patterns and changes in clinical scope but provided limited information on how patient demographics and referral indications were associated with whether genetic testing was ordered, which tests were used, and the likelihood of a diagnostic result. Such information is needed to inform adult referral and testing recommendations and to help clinicians counsel patients about expected diagnostic yield.

Understanding these patterns is particularly important when specialist capacity is limited and access is unequal. Reports of genetics service delivery describe long wait times, geographic barriers, and staffing constraints,^23,24^ while studies of adults show that Black patients and patients from economically disadvantaged neighborhoods are less likely to undergo genetics evaluation.^25^ Workforce surveys further document shortages and geographic concentration of genetics providers, with limited representation of physicians trained in internal medicine.^22,26,27^ Practice-based evidence linking patient characteristics, testing utilization, and outcomes is therefore needed to inform referral prioritization, guide test selection, and support planning for adult genetics services.

Herein, across 9,867 patient encounters spanning eight years, we characterize the clinical practice of adult general genetics from patient presentation and testing decisions through diagnostic outcomes and care delivery. By linking patient demographics and referral indications with testing utilization, diagnostic yield, and provider models, our study provides a practice-level view of how genomic medicine is implemented in adult patients. Our results demonstrate substantial diagnostic yield throughout adulthood, identify important differences in testing utilization and outcomes across indications and modalities, and highlight opportunities to develop more systematic pathways for referral, test selection, and service delivery.

## Subjects, Materials, and Methods

### Study population definition and data sources

This study was approved by the University of Pennsylvania’s Institutional Review Board (protocol #850058). We obtained from the Epic Clarity database the complete list of all 9,867 patient visits at the PennMedicine Perelman Center for Advanced Medicine’s Adult Genetics Clinic over an eight-year period from 10/1/2016 to 9/30/2024, along with accompanying visit demographic information including patient age, EHR-reported sex, race, ethnicity, home address, and financial class for the visit. Routine hereditary cancer, cardiac genetics, and neurogenetics evaluations are largely performed in separate PennMedicine specialty clinics, limiting the representation of these indications in this general adult genetics cohort. This information was joined with the internal clinical databases maintained by the PennMedicine Adult Genetics Clinic documenting visit indications, whether genetic testing was sent, and results of genetic testing. After joining, 4,264 patient visits had at least one missing data field, all of which were resolved by manual chart review by genetic counselors or physicians. Clinical indications for patient visits were collapsed into 21 broad indication groups (**Table S1**) by a panel of genetics providers blinded to both patient demographic information and outcomes of genetic testing. Latitudes and longitudes were obtained from patient home addresses using the tool Geocodio.

### Genetic Variant Classification

Genetic test results were classified as “pathogenic” if the variant identified was classified as pathogenic/likely pathogenic by the testing lab – for autosomal recessive conditions, two variants in the same gene were required with at least one classified as pathogenic/likely pathogenic – and was consistent with the patient’s presentation as determined by the clinical team at the time of result disclosure. Results were classified as “carrier” if a single pathogenic/likely pathogenic variant was identified in a gene associated with only autosomal recessive disease and no other pathogenic result had been reported. Results were classified as “candidate gene” if the testing lab had reported out only a variant in a candidate gene, i.e., a gene not definitively known to be associated with human disease, but suspicious enough to warrant reporting. Results were classified as variants of uncertain significance (VUS) if the lab reported only VUS(s). Lastly, genetic test reports that identified only likely-benign/benign variants or identified no causative variants, or exome sequencing (WES) and genome sequencing (WGS) results revealing only a secondary finding, were considered negative. Variant classification at the time of result disclosure to the patient was used throughout the study; thus, variants that subsequently changed classification on reanalysis were not updated for the sake of the analyses presented.

### Definitions of race and ethnicity

Race and ethnicity of the study population were extracted as reported in the EHR. These data likely represent a combination of self-reported and clinician/staff-assigned identities. Prior studies suggest that, although inaccuracies exist, EHR-derived race and ethnicity is highly concordant with self-reported data.^28^ Race and ethnicity represent distinct demographic identifiers; however, due to small sample sizes, we chose to combine race and ethnicity identifiers as follows. Any patient with an EHR-derived ethnicity of “Hispanic or Latino,” regardless of race, was collapsed into a single category, “Hispanic or Latino of any race” (n=339). For all patients without an EHR-derived ethnicity of “Hispanic or Latino,” race/ethnicity categories were defined as follows. Patients with an EHR-derived race listed as “other” (n=136), “some other race” (n=106), “unknown” (n=780), or “patient declined” (n=57) were listed as “Unknown/Other” (n=1,079). Patients with an EHR-derived race listed as “Black” (n=396) or “Black or African American” (n=349) were listed as “Black or African American” (n=745). Patients with an EHR-derived race listed as “American Indian” (n=6) or “American Indian or Alaskan Native” (n=12) were listed as “American Indian or Alaskan Native” (n=18). Patients with an EHR-derived race listed as “Pacific Island” (n=4) or “Native Hawaiian or Other Pacific Islander” (n=2) were listed as “Native Hawaiian or Other Pacific Islander” (n=6). Patients with an EHR-derived race listed as “Asian” (n=266) or “East Indian” (n=20) were listed as “Asian” (n=286). Patients with an EHR-derived race listed as “White” (n=7,384) were listed as “White” (n=7,384).

### Statistical analysis

Statistical analyses were carried out, as indicated, adjusting for covariates as described in the figure and table legends. For all analyses, two-sided p-values less than 0.05 were considered nominally significant. For analyses involving multiple hypothesis tests, statistical significance was determined using Bonferroni-adjusted p-values, with correction applied within the prespecified family of comparisons for each analysis, as detailed in the corresponding figure legends. Raw, unadjusted p-values are reported in the text, and Bonferroni-adjusted p-values are provided in the accompanying online analysis tables. All analyses were performed with R software, version 4.2.2.^29^

## Results

### Demographics of the PennMedicine Adult Genetics Clinic

We first evaluated geographic and demographic characteristics of the 9,867 unique patient visits for 7,766 unique patients to the PennMedicine Adult Genetics Clinic between October 1, 2016, and September 30, 2024. Most patients originated from the greater Philadelphia metropolitan area (**Figure 1A**). Although most patients clustered in or near Philadelphia, we observed substantial reach into surrounding regions, including parts of New Jersey, Delaware, and southeastern Pennsylvania. A smaller but notable number of patients traveled from further locations such as Baltimore, Washington D.C., and New York City, highlighting the clinic’s regional draw. Patients ranged widely in age at time of visit, from adolescence to older adulthood, but the overall distribution skewed younger, with the highest density of visits occurring among adults under 50 years of age (**Figure 1B**). Two distinct peaks were observed: one around age 30 years and another around age 60 years, in both male and female patients, suggesting different referral patterns or clinical needs across the age spectrum.

**Figure 1.**
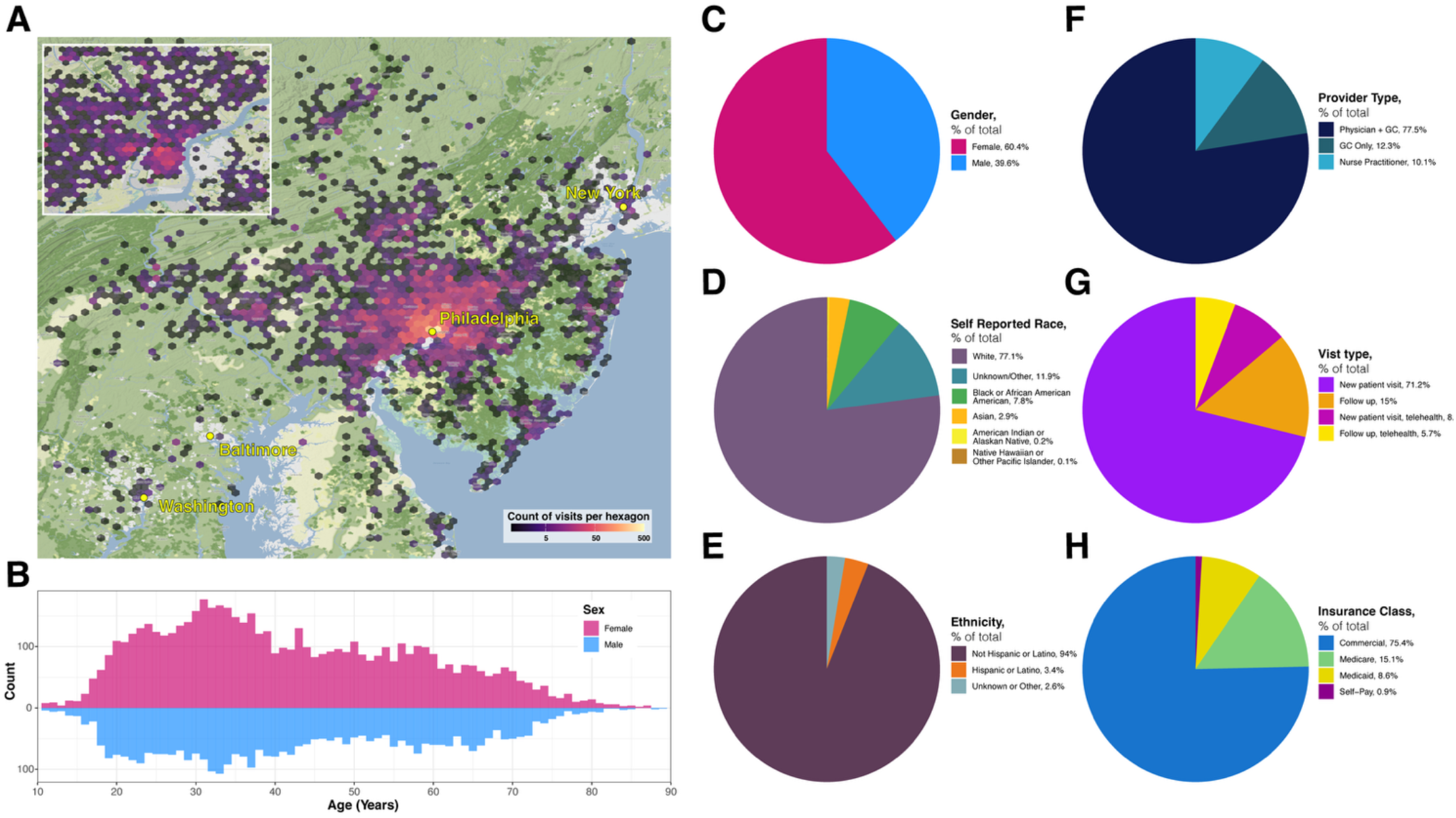
Demographics of the PennMedicine Adult Genetics Clinic. Demographics information for all 9,867 patient visits to the PennMedicine Adult Genetics Clinic are shown for the time period from October 1, 2016, to September 30, 2024. (A) A heatmap of patient home addresses is displayed over a geographic map of the Delaware Valley region, with major U.S. cities labeled for orientation. The inset provides a magnified view of the City of Philadelphia. The color scale indicates the number of unique patient visits per hexagonal unit. (B) Histogram illustrating the distribution of patient ages at time of visit, divided by EHR-reported sex. (C) EHR-reported patient sex for the study cohort. (D) EHR-reported patient race for the study cohort defined as reported in Subjects, Materials, and Methods. (E) EHR-reported patient ethnicity for the study cohort defined as reported in Subjects, Materials, and Methods. (F) Percent of visits conducted by each provider type across the entire study cohort (G) Visit type, divided by new vs. follow up visit, and by in-person and telehealth, for each patient visit. (H) EHR-reported patient insurance class at time of visit for the study cohort.

The clinic saw more female than male patients overall (60.4% vs. 39.6%) (**Figure 1C**). Most patients were reported in the EHR as White (77.1%), with the next most frequent groups being “Unknown/Other” (11.9%) and Black or African American (7.8%) (**Figure 1D**). Hispanic or Latino patients of any race comprised 3.4% of visits (**Figure 1E**). These proportions are consistent with known racial disparities in access to genetic services, as we have previously reported,^25^ and highlight an ongoing need for improved outreach and equity in adult genetics care.

The vast majority of patient visits were conducted by a genetics-trained physician and genetic counselor (77.5%), with 12.3% conducted by a genetic counselor alone and 10.1% (primarily for follow-up patient care) conducted by a nurse practitioner (**Figure 1F**). Most visits were for new patient consultations, either in person or via telehealth, together accounting for 79.3% of all encounters (**Figure 1G**). In-person and telehealth follow-up visits comprised the remaining 20.7% of encounters. Telehealth accounted for 13.8% of all visits, with 8.1% of all visits being for new consultations by telehealth and 5.7% for follow-up by telehealth (**Figure 1G**). A sharp rise in telehealth new patient and follow-up visits was observed beginning in 2020, coinciding with the onset of the COVID-19 pandemic and local stay-at-home orders (**Figure S1**).^30^ Although telehealth use declined in the subsequent years, a substantial proportion of follow-up visits continued to be conducted via telehealth (telehealth accounted for 38.8% of all follow-up visits from 2023 onwards), highlighting its enduring role in the delivery of adult genetics care for established patients. Most patients were commercially insured (75.4%), followed by those with Medicare (15.1%) and Medicaid (8.6%) (**Figure 1H**). Only 0.9% of visits were self-pay.

### Distribution and temporal trends in indications for clinic visits

Patients were referred to the PennMedicine Adult Genetics Clinic for a diverse array of clinical indications (**Figure 2A, S2**). The most common categories included cancer genetics (27.3%), connective tissue disorders (specifically inherited, non-immune connective tissue disorders, 16.6%), multisystem disorders (10.7%), joint hypermobility (11.8%), lysosomal storage disorders (5.8%), and vascular malformation disorders (4.6%). For cancer genetics specifically, patients are seen across multiple clinics at PennMedicine, with our program predominantly seeing patients with renal, neuroendocrine, skin, thyroid, brain and nerve sheath tumors. Referrals for hypermobility declined sharply and significantly over the study period (p<1e-04; OR 0.77 [0.76-0.79]), beginning in late 2019 (**Figure 2B**). This trend aligns with the implementation of new criteria for evaluation in our clinic in early 2020 which aimed to prioritize patients with more complex or multisystem presentations and reduce volume of lower-yield isolated hypermobility evaluations, for which there is no known genetic cause.^31^ As hypermobility referrals decreased, the clinic was able to accommodate patients with a broader range of referral indications. Resultant yearly increases were observed in referrals for cancer genetics (p<1e-04; OR 1.08 [1.07-1.09]), renal disorders (p<1e-04; OR 1.08 [1.05-1.11]), lung disorders (p<1e-04; OR 1.09 [1.05-1.13]), chromosomal disorders (p<1e-04; OR 1.06 [1.03-1.09]), lysosomal storage disorders (p=1.2e-04; OR 1.04 [1.02-1.06]), gastrointestinal disorders (p=1.52e-03; OR 1.13 [1.05-1.22]) and endocrine disorders (p=2.16e-03; OR 1.05 [1.02-1.08]) during this time period. These annual increases, accumulating over the eight-year study period, suggest a broadening of the clinic’s scope to serve patients with a more diverse range of conditions requiring genetic evaluation.

**Figure 2.**
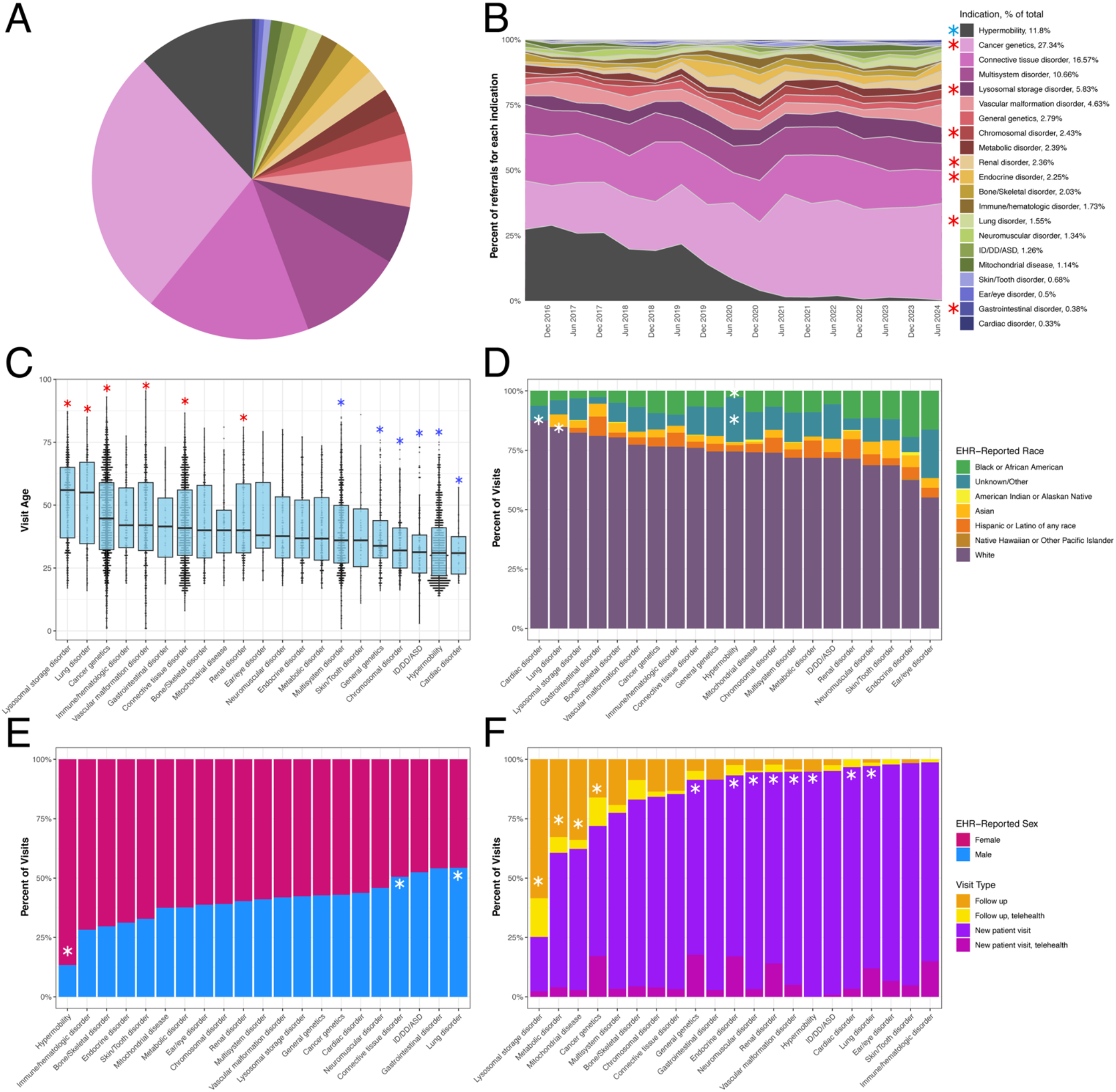
Comparison of Demographic Characteristics and Visit Types Across Clinical Indications. (A) Indications for all 9,867 unique patient visits to the PennMedcine Adult Genetics Clinic from October 1, 2016, to September 30, 2024 are displayed. The percentage of each indication, relative to the total, is shown in the legend “Indication Group, Percent of Total”. (B) Patient visit indications are shown over time in 6-month intervals from October 1, 2016, to September 30, 2024. Indications are color-coded as shown in the legend “Indication Group, Percent of Total.” Indications with a significant decrease in their proportion over time (Hypermobility) are marked with a blue asterisk in the legend, while those with a significant increase (Cancer genetics, Lysosomal storage, Chromosomal disorder, Renal disorder, Endocrine disorder, Lung disorder, Gastrointestinal disorder) are marked with a red asterisk. Statistical significance was determined by logistic regression accounting for multiple testing across all 21 indication groups. (C) Dot plots with overlaid boxplots show patient age at the time of visit for each indication group, ordered by median age, across the study period. Each dot represents an individual patient, while boxplots display the median (solid line) and interquartile range (box and whiskers). Indications with a mean patient age significantly above the clinic-wide average (Lysosomal storage disorder, Lung disorder, Cancer genetics, Vascular malformation, Connective tissue disorder, Renal disorder) are marked with red asterisks; those significantly below (Cardiac disorders, Hypermobility, ID/DD/ASD, Chromosomal disorder, General genetics, Multisystem disorder) are marked with blue asterisks. Statistical significance was determined by linear regression adjusting for EHR-reported sex, race, insurance class, and visit type, with Bonferroni correction applied for multiple testing across all 21 indication groups. (D) Stacked bar charts display the EHR-reported race of patients for each indication group across the study period. Indications with a significantly higher proportion of a given racial group, compared to the overall clinic average, are marked with an asterisk. Statistical significance was determined by multinomial logistic regression, adjusting for sex, insurance class, visit type, and age at visit. After Bonferroni correction, patients of Unknown/Other race were found to be significantly overrepresented among referrals for Hypermobility, while patients of Black or African American race were significantly underrepresented. Hispanic or Latino patients were underrepresented in referrals for cardiac and lung disorders. Patients of Black or African American race were significantly overrepresented in referrals for Endocrine disorders and Cancer genetics. (E) Stacked bar charts display the proportion of male and female patients for each indication group across the study period. Indications with a significantly higher proportion of female patients compared to the overall clinic average (Hypermobility) are marked with an asterisk in the pink bar, while those with a significantly higher proportion of male patients compared to the overall clinic average (Connective tissue disorder, Lung disorder) are marked with an asterisk in the blue bar. Statistical significance was determined by logistic regression, adjusting for age, race, insurance class, and visit type, with Bonferroni correction applied for multiple testing across all 21 indication groups. (F) Stacked bar charts display the proportion of new patient visits and follow-up visits, further subdivided by in-person and telemedicine encounters, for each indication group across all unique patient visits during the study period. Indications with a significantly higher proportion of follow up visits (in-person or telemedicine) compared to the overall clinic average (Lysosomal storage disorder, Cancer genetics, Metabolic disorder, Mitochondrial disease), are marked with an asterisk in the orange bar, while those with a significantly higher proportion of new patient visits (in-person or telemedicine) compared to the overall clinic average (General genetics, Endocrine disorder, Neuromuscular disorder, Renal disorder, Vascular malformation disorder, Hypermobility, Cardiac disorder, Lung disorder) are marked with an asterisk in the purple bar. Statistical significance was determined by logistic regression, adjusting for EHR-reported sex, race, insurance class, and age at visit, with Bonferroni correction applied for multiple testing across all 21 indication groups.

Within these broad indication categories, the most common specific indications driving clinic visits (**Figure S2**) were hypermobility of the joints (1,157 visits, 11.8% of all visits), followed by von Hippel-Lindau disease (576 visits, 5.9%), Neurofibromatosis type 1 (NF1; 499 visits, 5.1%), aneurysmal disease (427 visits, 4.4%), Marfan syndrome (424 visits, 4.3%), Hereditary Hemorrhagic Telangiectasia (374 visits, 3.8%), Fabry disease (315 visits, 3.2%), *SDHB*-related cancer predisposition syndrome (228 visits, 2.3%), Tuberous Sclerosis Complex (TSC; 186 visits, 1.9%), and arterial dissection (181 visits, 1.9%). Together, these ten indications accounted for 44.3% of all clinic visits, reflecting the concentration of referrals around a relatively small number of well-defined heritable conditions. The preponderance of connective tissue and vascular diagnoses – including Marfan syndrome, aneurysmal disease, arterial dissection, and hereditary hemorrhagic telangiectasia – underscores the clinic’s established role as a referral center for adults with heritable aortopathy and related vascular disorders. The prominence of multisystem tumor predisposition syndromes, including von Hippel-Lindau, NF1, TSC, and SDHB-related cancer predisposition, reflects the clinic’s substantial engagement with hereditary tumor syndrome management alongside traditional cancer genetics referrals.

### Significant variation in patient age, race, and sex across clinical indication groups

Significant demographic differences were observed among patients referred for different clinical indications (**Figure 2C–F**). Patients referred for lysosomal storage disorders and lung disorders had the highest median ages (56 and 55 years, respectively), whereas patients referred for hypermobility and cardiac disorders had the lowest (31 and 30 years, respectively, **Figure 2C**). After adjusting for EHR-reported sex, race, insurance class, and visit type, patients referred for cancer genetics (p<1e-04; beta 4.36 [3.22,5.49]), connective tissue disorders (p<1e-04; beta 2.09 [0.81,3.38]), lung disorders (p<1e-04; beta 5.37 [1.99,8.76]), lysosomal storage disorders (p<1e-04; beta 10.32 [8.35,12.29]), renal disorders (p=7.7e-04; beta 3.21 [0.45, 5.96]), and vascular malformation disorders (6.8e-04; beta 2.42 [0.39, 4.46]) were significantly older than the mean age of overall clinic population (39 years). In contrast, patients referred for cardiac disorders (p=1.2e-03; beta -8.03 [-15.21, -0.86]), chromosomal disorders (p<1e-04; beta -5.87 [-8.58,-3.16]), general genetics (1.7e-04; beta -3.94 [-6.47,-1.42]), hypermobility (p<1e-04; beta -6.48 [-7.95,-5.00]), ID/DD/ASD (Intellectual Disability/Developmental Delay/Autism Spectrum Disorders, p<1e-04; beta -9.14 [-12.80,-5.49]), and multisystem disorders (p<1e-04; beta -1.73 [-3.21, - 0.25]) were significantly younger than mean age of the overall clinic population, after correcting for multiple comparisons.

Differences in EHR-reported race and ethnicity across clinical indication groups were modest in magnitude, although several remained statistically significant after covariate adjustment and correction for multiple comparisons. Examining EHR-reported patient race by indication group (**Figure 2D**), we found that patients evaluated for hypermobility were significantly more likely to have an EHR-reported race of Unknown/Other (p<1e-04; OR 1.09[1.06-1.13]) and significantly less likely to have an EHR-reported race of Black or African American (p<1e-04; OR 0.95 [0.93-0.96]), by logistic regression after adjusting for EHR-reported sex, age, insurance class, visit type, and after correcting for multiple comparisons. Hispanic or Latino patients were significantly less likely to be evaluated for cardiac disorders (p=4.1e-04; OR 0.92[0.89-0.96]) and lung disorders (p=4.1e-04; OR 0.92[0.89-0.96]), whereas patients of Black or African American race were significantly more likely to be evaluated for endocrine disorders (p=2.0-03; OR 1.09[1.04-1.14]) and cancer genetics (p=2.3-03; OR 1.03[1.01-1.05]).

Significant differences in EHR-reported patient sex were also identified by indication group (**Figure 2E**). Patients referred for connective tissue disorders (p<1e-04; OR 1.52 [1.34–1.72]) and lung disorders (p=1.2e-03; OR 1.87 [1.35–2.59]) were significantly more likely to be male, whereas those referred for hypermobility were significantly less likely to be male (p<1e-04; OR 0.24 [0.20–0.28]), by logistic regression after adjusting for EHR-reported race, age, insurance class, visit type, and correcting for multiple comparisons.

Comparing visit type by indication group (**Figure 2F**) by logistic regression, adjusted for EHR-reported sex, race, age, and insurance class, we found that patients seen for certain clinical indications were significantly more likely to be seen for follow up visits (across both in person and telehealth visits) compared to the overall clinic population: lysosomal storage disorder (p<1e-04; OR 1.80 [1.67-1.93]), cancer genetics (p=3.3e-04; OR 1.13[1.07-1.20), metabolic disorder (p<1e-04; OR 1.24[1.14-1.36]), and mitochondrial disease (p=7.1e-04; OR 1.23[1.11-1.35]). On the other hand, patients seen for lung disorders (p<1e-04; OR 1.11[1.07-1.16]), cardiac disorders (p=1.49e-03; OR 1.12[1.05-1.18]), hypermobility (p<1e-04; OR 1.10[1.07-1.13]), vascular malformation disorders (p<1e-04; OR 1.09[1.06-1.12]), renal disorders (p<1e-04; OR 1.09[1.06-1.13]), neuromuscular disorders (p=1.0e-03; OR 1.09[1.04-1.13]), endocrine disorders (p=5.78e-04; OR 1.08[1.04-1.12]), and general genetics (p=8.2e-04; OR 1.06[1.03-1.10]) were significantly more likely to be seen for new patient visits (both in person and telehealth) compared to the overall clinic population.

### Higher genetic testing utilization among older patients and male patients

We next compared the likelihood of genetic testing being sent following evaluation. Overall, we found that genetic testing was completed in 51.9% of all new patients, and in 8.0% of all follow up patients during the study period (**Figure 3A**). Among new patient visits, male patients were significantly more likely than females to undergo genetic testing (57.2% of male tested compared to 45.8% of females, p=5.2e-03; OR 1.16[1.05-1.29]), even after adjusting for EHR-reported race, age, insurance class, and clinical indication (**Figure 3B**). This finding remained significant even when removing hypermobility evaluations from the analysis (**Figure S1E**). We also found a significant positive correlation between patient age at time of new patient evaluation and likelihood of completing genetic testing (p=1.2e-14; OR 1.01[1.01-1.02], with the OR reflecting the per-year change in odds of undergoing genetic testing after evaluation), even after adjusting for EHR-reported race, sex, insurance class, and clinical indication (**Figure 3C**). This finding remained significant even when removing hypermobility evaluations from the analysis (**Figure S1F**). No significant differences were seen in genetic testing rates across different race/ancestry groups after adjusting for multiple hypothesis testing (**Figure 3D**).

**Figure 3.**
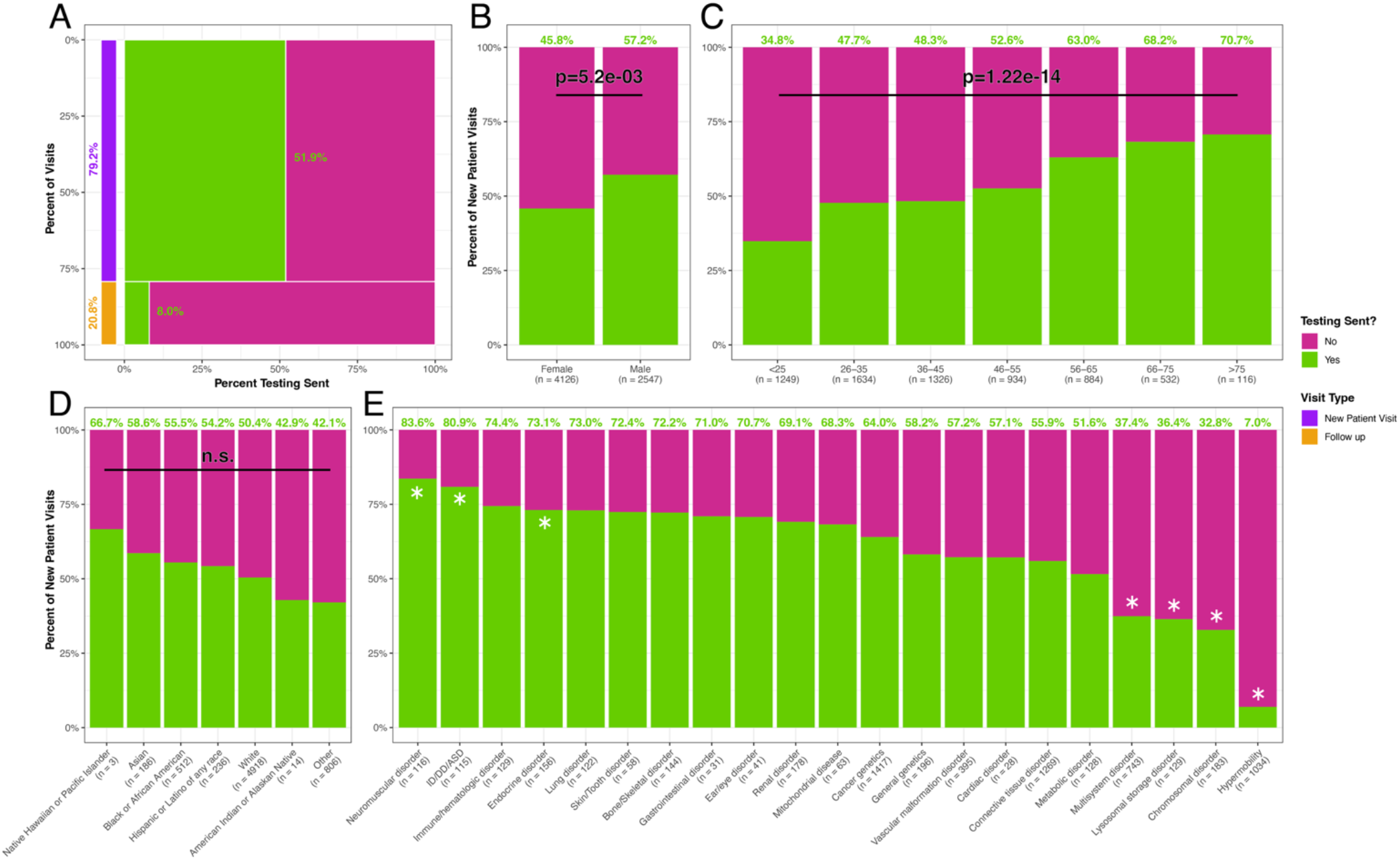
Patterns of Genetic Testing Utilization by Visit Type, Demographics, and Clinical Indication. In all panels, bars indicate the proportion of visits for which testing was sent (green) versus not sent (pink). Percent labels, in green, above bars show the percent of visits for which testing was sent for each subgroup. Sample sizes for each subgroup are shown below the horizontal axis labels for panels B-E. (A) Marimekko plot showing the percent of visits during which genetic testing was sent or not sent by visit type (New Patient Visit vs. Follow-up). The height of each row represents the proportion of all visits accounted for by each visit type; the width of each column represents the percent of visits, within each visit type, with or without testing sent. A stacked bar plot to the left shows the overall distribution of visit types, for reference, with New Patient Visits in purple and Follow-up visits in orange. (B) Percent of new patient visits during which testing was sent, stratified by EHR-reported sex. The p-value indicates the result of logistic regression comparing the odds of testing sent by sex, adjusted for clinical indication group, age, race, and insurance class. (C) Percent of new patient visits during which testing was sent, stratified by patient age (plotted in discrete age ranges). The p-value indicates the association between age (modeled as a continuous variable) and the odds of testing being sent, from logistic regression adjusted for clinical indication group, race, sex, and insurance class. (D) Percent of new patient visits during which testing was sent, stratified by EHR-reported race. No significant differences were found when comparing each group to the largest group (EHR-reported White race) by logistic regression adjusted for indication group, sex, age, and insurance class, accounting for multiple testing across all 6 groups. (E) Percent of new patient visits during which testing was sent, stratified by clinical indication group. Indications with a significantly higher odds of testing compared to the overall clinic average (Neuromuscular disorder, ID/DD/ASD, and endocrine disorder) are marked with an asterisk in the green bar, while those with a significantly lower odds of testing compared to the overall clinic average (Multisystem disorder, Lysosomal storage disorder, Chromosomal disorder, Hypermobility) are marked with an asterisk in the pink bar. Statistical significance was determined by logistic regression, adjusting for age, race, sex, and insurance class, with Bonferroni correction applied for multiple testing across all 21 indication groups.

### Significant differences in genetic testing utilization by clinical indication group

Significant differences were seen across referral indications for the propensity to complete genetic testing after new-patient evaluation (logistic regression adjusted for age, race/ethnicity, sex, and insurance class, **Figure 3E**). Relative to the clinic-wide new-visit average, testing was more likely to be ordered and completed for neuromuscular disorders (83.6% of new patients completed testing, p<1e-04; OR 3.13[1.98-4.96]), ID/DD/ASD (80.9%, p<1e-04; OR 2.71[1.73-4.25]), and endocrine disorders (73.1%, p=1.3e-03; OR 1.77[1.29-2.43]). In contrast, testing was less likely to be ordered for hypermobility (7.0%, p<1e-04; OR 0.06[0.04-0.07]), chromosomal disorders (32.8%, p<1e-04; OR 0.36[0.27-0.49]), lysosomal storage disorders (36.4%, p<1e-04; OR 0.39[0.28-0.55]), and multisystem disorders (37.4%, p<1e-04; OR 0.42[0.35-0.49]). Our data do not allow us to assess whether lower genetic test ordering rates in these groups reflect prior confirmatory testing obtained before referral to our clinic. We suspect that many new patients being referred for chromosomal disorders, lysosomal storage disorders, or known multisystem disorders already carried a molecular diagnosis and testing was not required.

### Genetic testing modalities vary by indication and over time

Patterns of genetic testing modalities and laboratory utilization shifted during the study period (**Figure 4A-B**). Of the 4,245 tests sent during this period, the most common type of test sent was multi-gene panels (NGS, 62.84%), followed by single gene (15.23%), single site (10.80%), exome sequencing (3.69%), and chromosomal microarray (CMA, 2.68%). Over time, the ordering of multi-gene panels (p=4.2e-05; OR 1.03 [1.02–1.04]) and genome sequencing (WGS, p=3.2e-03; OR 1.35 [1.13–1.71]) increased significantly, whereas significant decreases were observed in the ordering of CMA (p=8.8e-07; OR 0.90 [0.86–0.94]), single site testing (p=1.5e-04; OR 0.96 [0.94–0.98]), and metabolic testing (i.e. non-genetic biochemical testing, p=1.4e-03; OR 0.92 [0.87–0.97]).

**Figure 4.**
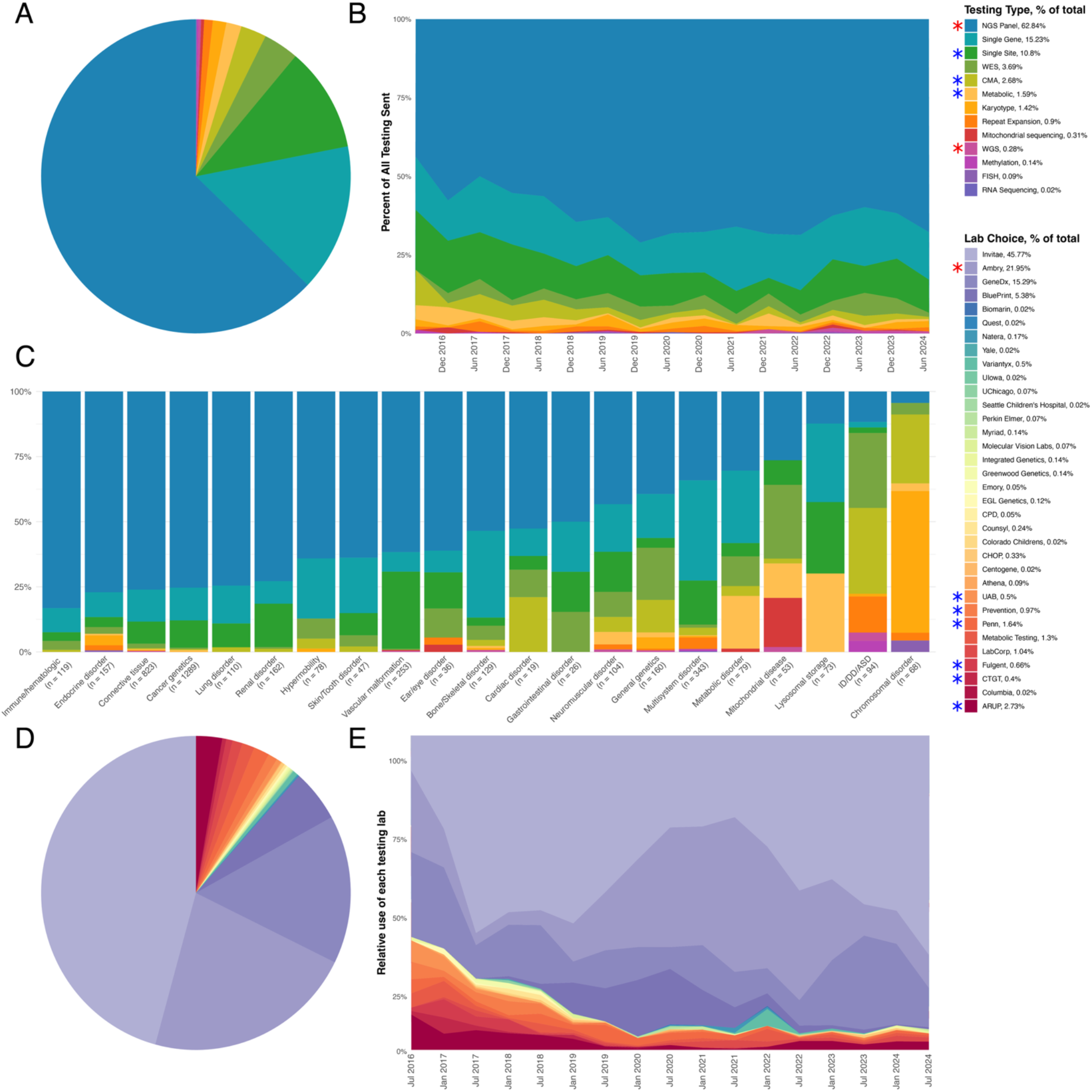
Patterns and Temporal Trends in Genetic Testing Modalities and Laboratory Utilization. (A) Type of testing sent for all 4,245 tests sent during the study period. Percentages relative to the total are shown in the legend “Testing Type, Percent of Total.” (B) Type of testing sent is shown over time in 6-month intervals from October 1, 2016, to September 30, 2024. Testing types are color-coded as shown in the legend “Testing Type, Percent of Total.” Testing types significantly increasing in utilization over time (NGS Panel, WGS) are marked with a red asterisk in the legend, while those significantly decreasing in utilization over time (Single Site, CMA, Metabolic) are marked with a blue asterisk. Statistical significance was determined using separate binary logistic regression models for each test type, comparing the odds of using that test type (versus all other tests) as a function of time, accounting for multiple testing across all 13 testing types. (C) Type of testing sent, by clinical indication group, for all tests sent during the study period. Each bar shows the proportion of tests of each type within an indication group, color-coded as in the legend “Testing Type, Percent of Total”; the number of total tests in each group is shown beneath the axis label. Indication groups are ordered from left to right by decreasing proportion of NGS panels. (D) Testing laboratories used for all 4,245 tests sent during the study period. Percentages relative to the total are shown in the legend “Lab Choice, Percent of Total.” (E) Testing lab choice is shown over time in 6-month intervals from October 1, 2016, to September 30, 2024. Testing labs are color-coded as shown in the legend “Lab Choice, Percent of Total.” Testing labs significantly increasing in utilization over time (Ambry) are marked with a red asterisk in the legend, while those significantly decreasing in utilization over time (UAB, Prevention, Penn, Fulgent, CTGT, ARUP) are marked with a blue asterisk. Statistical significance was determined using binary logistic separate regression models for each testing lab, comparing the odds of using that test type (versus all other labs) as a function of time, accounting for multiple testing across all 34 testing labs.

Test selection also varied substantially by clinical indication (**Figure 4C**). Multi-gene panels, single-gene tests, and single-site familial-variant tests together accounted for more than 95% of testing ordered for vascular malformation disorders (98.8%), cancer genetics (98.4%), lung disorders (98.2%), renal disorders (98.1%), connective tissue disorders (96.8%), and immune/hematologic disorders (95.8%). At the same time, these three modalities accounted for a much smaller proportion of testing for mitochondrial disease (35.8%), ID/DD/ASD (16.0%), and suspected chromosomal disorders (4.4%). For mitochondrial disease, the most frequently ordered tests were WES (28.3%), multi-gene panels (26.4%), and mitochondrial genome sequencing (18.9%). For ID/DD/ASD the most frequently ordered tests were CMA (33.0%), WES (28.7%), and repeat-expansion testing (13.8%). Testing for suspected chromosomal disorders consisted primarily of karyotyping (54.4%) and CMA (26.5%), with fluorescence in situ hybridization (FISH), WES, and NGS panels each accounting for 4.4%. For all other indication groups, NGS, single-gene tests, and single-site familial-variant tests accounted for between 60-95% of all testing sent.

### Visit types, testing, and test selection vary by provider type

Provider types differed in the visits they conducted and the testing they ordered (**Figure S1B-D**). Visits with a physician and genetic counselor were predominantly new-patient evaluations (83.3%), almost always in person (2.9% of new-patient visits by telehealth). Genetic counselor-only visits were likewise predominantly new-patient evaluations (95.0%), but 51.1% of these were conducted by telehealth, and counselor-only visits accounted for 73.4% of all new-patient telehealth visits in the clinic (**Figure S1A-B**). Nurse practitioner visits were predominantly follow-up (69.0%), 38.5% of which were by telehealth. Testing was sent at 80.9% of counselor-only new-patient visits, compared with 48.7% of physician-led and 12.1% of nurse practitioner new-patient visits (**Figure S1C**). Test selection differed by provider type as well (**Figure S1D**). At counselor-only visits, single-site familial-variant tests accounted for 26.2% of tests sent, compared with 6.5% at physician-led visits, whereas broad or specialized modalities (WES, WGS, CMA, karyotype, metabolic, and mitochondrial testing) accounted for 11.5% of tests sent at physician-led visits but only 2.4% at counselor-only visits. Multi-gene panels were the most common test type for all three provider types (56.9-67.6%).

### Choice of testing lab shifted alongside changes in cost, coverage, and ordering infrastructure

Testing was sent to 34 different laboratories (**Figure 4D**), with three laboratories performing 83.01% of all testing during the study period; 1,945 tests were performed at Invitae (45.82%), 931 were performed at Ambry Genetics (21.93%), and 648 were performed at GeneDx (15.27%). As with the types of tests ordered, temporal shifts were also observed in which genetic testing laboratories were used (**Figure 4E**). Over the study period, the ordering of testing from Ambry increased significantly (p=5.23e-08; OR 1.05 [1.03–1.06]), whereas significant decreases were observed in testing ordered from ARUP (p=3.19e-06; OR 0.90 [0.87–0.94]), CTGT (p=4.8e-06; OR 0.59 [0.46–0.72]), Fulgent (p=7.86e-09; OR 0.62 [0.52– 0.72]), Prevention (p=1.85e-09; OR 0.75 [0.68–0.82]), University of Alabama (UAB, p=1.39e-07; OR 0.49 [0.36–0.62]), and the University of Pennsylvania (Penn, p=3.88e-04; OR 0.91 [0.86–0.96]). No statistically significant differences were observed in the utilization of other laboratories during this time.

Year-to-year shifts in testing laboratories closely tracked changes in billing, insurance, and ordering policies (**Figure 4E**). For example, Blueprint Genetics introduced a $250 capped-cost policy for self-pay NGS panels in November 2018, which led to a sharp increase in orders, but discontinued the policy in February 2021, after which orders declined. Variantyx offered no-cost reflex from any NGS panel to genome sequencing between January 2022 and February 2023, producing a similar rise and fall in orders during that period. Test ordering for Ambry was integrated into the PennMedicine electronic medical record in January 2020, resulting in an increase in testing volume, and GeneDx became in-network with Independence Blue Cross in February 2023, likewise driving an increase in tests ordered.

### High rate of positive findings across all patients

Overall, a high rate of pathogenic findings was observed across all testing sent during the study period (**Figure 5A**). Of the 4,245 tests sent, 24.3% resulted in a pathogenic, diagnostic finding. Another 3.8% returned pathogenic carrier results, and 13.6% returned inconclusive (VUS) results.

**Figure 5.**
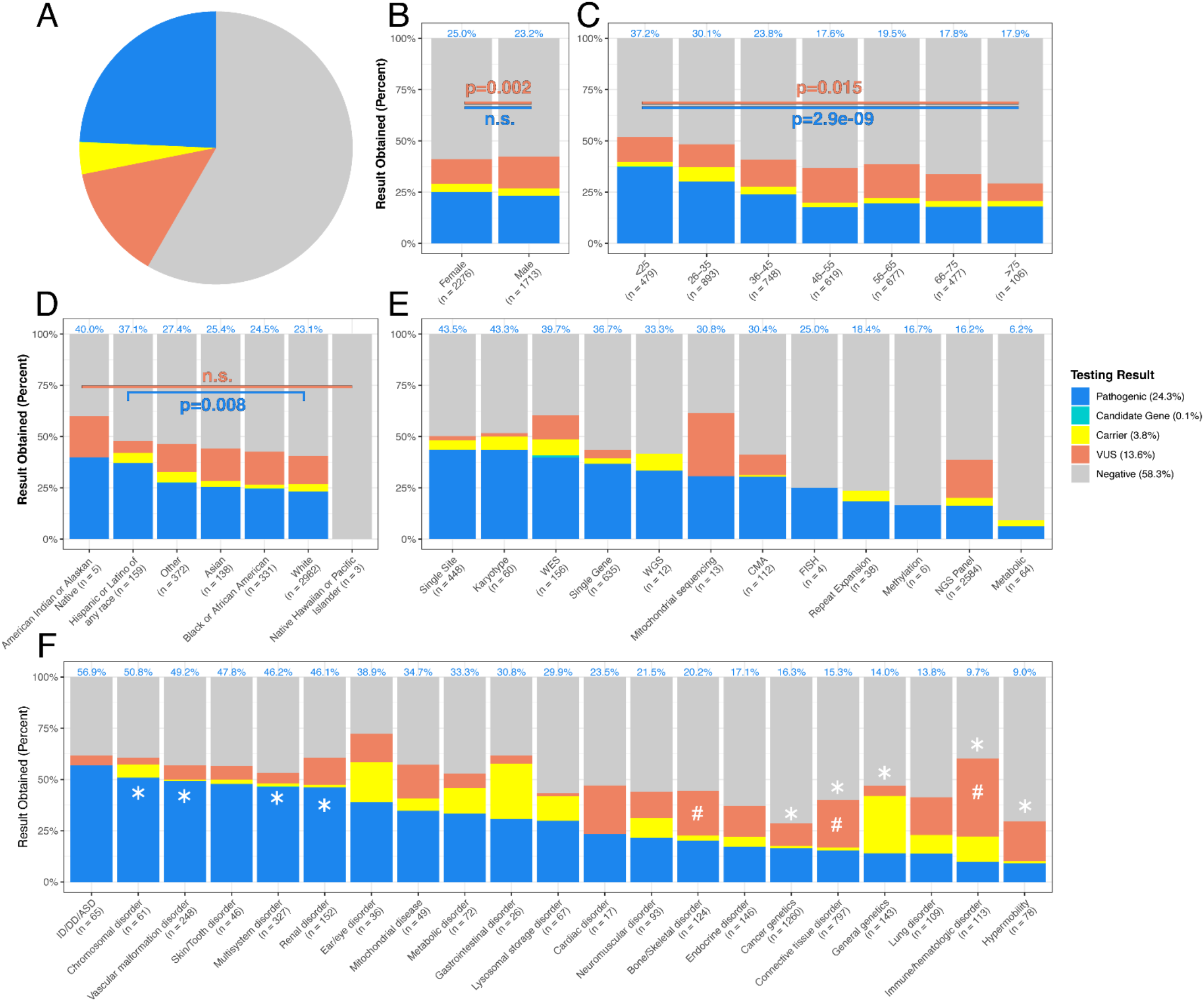
Genetic Testing Results by Patient and Test Characteristics. In all panels, bars indicate the percentage of patients with each genetic testing result classification: pathogenic (blue), variant of uncertain significance (VUS; orange), carrier (yellow), candidate gene (aqua, present only in WES and WGS testing), or negative (gray). Percent labels in blue above bars show the percent of patients who received a pathogenic result for each subgroup. Sample sizes for each subgroup are shown below the horizontal axis labels for panels B–F. (A) Pie chart showing the overall distribution of results for all 4,245 genetic tests sent during the study period. Percentages for each result classification are shown in the legend. (B) Percent of patients receiving each testing result classification, stratified by EHR-reported sex. The p-values indicate the result of logistic regression comparing the odds of receiving a pathogenic (in blue) or VUS (in orange) result by sex, adjusted for patient age, race, insurance class, clinical indication group, and visit type. No significant difference was found for pathogenic results; however, males were significantly more likely than females to receive a VUS result. (C) Percent of patients receiving each testing result classification, stratified by patient age at visit (plotted in discrete age ranges). The p-value indicates the association between age (modeled as a continuous variable) and the odds of receiving a pathogenic (in blue) or VUS (in orange) result from logistic regression adjusted for sex, race, insurance class, clinical indication group, and visit type. Older patients were significantly less likely to receive a pathogenic result and significantly more likely to receive a VUS. (D) Percent of patients receiving each testing result classification, stratified by EHR-reported race. Hispanic or Latino patients of any race were significantly more likely to receive pathogenic results compared to patients of EHR-reported White race; no other significant differences were found for any racial group, relative to patients with EHR-reported White race, for the odds of receiving either a pathogenic (in blue) or VUS (in orange) result by logistic regression adjusted for age, sex, insurance class, clinical indication group, and visit type, accounting for multiple testing across all 6 groups. (E) Percent of patients receiving each testing result classification, stratified by type of testing, ordered by likelihood of receiving a pathogenic result. No formal statistical comparisons were performed for this panel. (F) Percent of patients receiving each testing result classification, stratified by clinical indication group, ordered by likelihood of receiving a pathogenic result. Indications where genetic testing had a significantly higher odds of identifying a pathogenic result compared to the overall clinic average (Chromosomal disorder, Vascular malformation disorder, Renal disorder, Multisystem disorder) are marked with an asterisk in the blue bar, while those with a significantly lower odds of identifying a pathogenic result compared to the overall clinic average (Cancer genetics, Connective tissue disorder, General genetics, Immune/hematologic disorder, Hypermobility) are marked with an asterisk in the grey bar. Indications where genetic testing had a significantly higher odds of identifying a VUS compared to the overall clinic average (Connective tissue disorder, Immune/hematologic disorder, bone/skeletal disorders) are marked with a hash mark in the orange bar. Statistical significance was determined by logistic regression, adjusting for age, race, sex, and insurance class, with Bonferroni correction applied for multiple testing across all 21 indication groups.

The percentage of patients receiving each testing result was stratified by EHR-reported sex (**Figure 5B**). The rate of pathogenic findings did not vary significantly between EHR-reported sex; 25.0% of females and 23.2% of males received a pathogenic result. However, males were significantly more likely than females to receive a VUS result, with 15.6% of testing sent in male patients and 12.0% of testing sent in female patients resulting with one or more VUSs (p=0.002; OR 1.34 [1.11-1.61], results of logistic regression adjusted for age, race, insurance class, clinical indication group, and visit type).

The percentage of patients receiving each testing result was also stratified by EHR-reported race and ethnicity (**Figure 5D**). The rate of pathogenic findings was 40.0% for American Indian or Alaskan Native patients (n=5), 38.1% among Hispanic or Latino patients (n=155), 27.4% for patients of Unknown/Other race/ethnicity (n=372), 25.4% for Asian patients (n=138), 24.5% for Black or African American patients (n=331), 23.1% for White patients (n=2,986), and 0% for Native Hawaiian or Other Pacific Islander patients (n=3). Hispanic or Latino of any race were significantly more likely to receive pathogenic results than White patients (p=1.32e-03, OR 1.81 [1.25-2.59]) by logistic regression adjusted for age, sex, insurance class, clinical indication group, and visit type; no other significant differences in pathogenic result rate were observed by EHR-reported race/ethnicity. The rate of VUS findings was 20.0% for American Indian or Alaskan Native patients, 5.7% among Hispanic or Latino patients, 13.9% for patients of Unknown/Other race/ethnicity, 15.8% for Asian patients, 16.4% for Black or African American patients, 13.7% for White patients, and 0% for Native Hawaiian or Other Pacific Islander patients. No significant differences were observed for VUS rate across patients of different racial/ethnic groups.

### Significant decrease in positive testing rate with increasing patient age

When analyzing results of genetic testing by patient age, the percentage of patients receiving each testing result classification differed significantly (**Figure 5C**). Older patients were significantly less likely to receive a pathogenic result overall (p=1.80e-09; OR 0.98[0.98-0.99], results of logistic regression adjusted for race, sex, insurance class, clinical indication group, and visit type, OR reflects the per-year-of-age change in odds of having a pathogenic result). Pathogenic results were identified in 37.2% of patients aged 18-25 years (n=479), 30.1% of patients 26-35 years (n=893), 23.8% of patients 36-45 years (n=748), 17.6% of patients 46-55 years (n=619), 19.5% of patients 56-65 years (n=677), 17.8% of patients 66-75 years (n=477), and 17.9% of patients >75 years (n=106). Older patients were also nominally significantly more likely to receive a VUS result (p=6.26e-03; OR 1.01[1.00-1.02], results of logistic regression adjusted for age, sex, insurance class, clinical indication group, and visit type, OR reflects the per-year change in odds). VUS results were identified in 11.9% of patients aged 18-25 years, 11.2% of patients 26-35 years, 13.0% of patients 36-45 years, 17.0% of patients 46-55 years, 16.7% of patients 56-65 years, 13% of patients 66-75 years, and 8.5% of patients >75 years.

### Variation in diagnostic yield across genetic testing modalities

Large differences in diagnostic yield were observed across different genetic testing modalities (**Figure 5E**). The testing modalities with the highest percentage of pathogenic findings were single site testing (43.5%, n=448), karyotype (43.3%, n=60), exome sequencing (WES, 39.7%, n = 156), single gene testing (36.7%, n=635), and chromosomal microarray (CMA, 30.4%, n=112). Next-generation sequencing (NGS) panels, by far the most common test ordered (n=2,584) had the second lowest diagnostic rate (16.2% diagnostic rate) of all testing modalities, followed only by metabolic testing (6.2% diagnostic rate, n=64). The rate of VUS results also varied between testing modalities. The testing types most likely to result in only VUSs were mitochondrial sequencing (30.8%), NGS panels (18.6%), exome sequencing (11.5%), and CMA (9.8%).

### Significant indication-specific variation in genetic test outcomes

The pathogenic-result rates varied more than six-fold across indication groups, from 9.0% in hypermobility to 56.9% in ID/DD/ASD. The highest rates were observed in ID/DD/ASD (56.9%, *n*=65), chromosomal disorders (50.8%, *n*=61), vascular malformation disorders (49.2%, *n*=248), skin/tooth disorders (47.8%, *n*=46), multisystem disorders (46.2%, *n*=327), and renal disorders (46.1%, *n*=152). The lowest rates occurred in hypermobility (9.0%, *n*=78), immune/hematologic disorders (9.7%, *n*=113), and lung disorders (13.8%, *n*=109). In covariate-adjusted comparisons, chromosomal (p=9.3e-04; OR 2.35 [1.46–3.78]), vascular malformation (p<1e-04; OR 2.87 [2.20–3.75]), renal (p<1e-04; OR 2.43 [1.76–3.35]), and multisystem referrals (p<1e-04; OR 2.26 [1.78–2.86]) had significantly higher odds of a pathogenic result than the overall modeled average, whereas cancer (p<1e-04; OR 0.64 [0.53–0.77]), connective tissue (p<1e-04; OR 0.55 [0.45–0.69]), general genetics (p=2.3e-04; OR 0.40 [0.26–0.63]), immune/hematologic (p=3.5e-04; OR 0.32 [0.18–0.57]), and hypermobility referrals (p=9.3e-04; OR 0.25 [0.12–0.54]) had lower odds after Bonferroni correction. VUS rates showed a different pattern, with the highest rates in immune/hematologic disorders (38.1%, *n*=113), cardiac disorders (23.5%, *n*=17), connective tissue disorders (23.0%, *n*=797), bone/skeletal disorders (21.8%, *n*=124), and hypermobility (19.2%, *n*=78). In adjusted analyses, connective tissue (p<1e-04; OR 2.48 [1.91–3.23]), immune/hematologic (p<1e-04; OR 5.80 [3.84–8.76]), and bone/skeletal referrals (p=8.1e-04; OR 2.45 [1.55–3.86]) had significantly higher odds of VUS results than the overall modeled average after Bonferroni correction.

Although testing outcomes varied across indications, so did the underlying demographic and clinical characteristics of the patients evaluated. Although earlier analyses demonstrated differences in age, sex, and race across indication groups (**Figure 2**), **Figure S3** provides a more detailed view of the particular clinical presentations represented at different patient ages within each indication group, together with their associated testing outcomes. Certain indication groups clearly had distinct clinical components driving genetic testing outcomes at different ages. For example, connective tissue referrals included a younger component enriched for Marfan syndrome and an older component enriched for aneurysmal disease; pathogenic-result density was more concentrated in the younger component (**Figure S3B**). Similarly, referrals for lung disease showed a peak among younger patients for pneumothorax and cystic fibrosis and a separate peak among older patients for interstitial lung disease and idiopathic pulmonary fibrosis; pathogenic-result density was again more concentrated among younger patients (**Figure S3M**). Together, these patterns indicate that indication-specific testing outcomes likely reflect both differences in diagnostic yield across indication groups and differences in the demographic and clinical composition of patients referred and tested for each indication.

### Genetic variants identified span a diverse set of genes

Across all 4,245 tests sent, variants in 514 unique single genes and sixty-nine unique chromosomal aneuploidies/abnormalities were identified (**Figure 6**). The most frequently identified single-gene variants were in *ACVRL1* (65 pathogenic variants and five VUSs identified), *FBN1* (38 pathogenic variants, 28 VUSs), *NF1* (57 pathogenic variants, seven VUSs), *ENG* (39 pathogenic variants, seven VUSs), *FLCN* (37 pathogenic variants, six VUSs), *SDHB* (39 pathogenic variants, four VUSs), and *PKD1* (26 pathogenic variants, 11 VUSs). The only recurrent chromosomal differences identified were 45,X in 16 individuals, 47,XXY in seven individuals, del(15)(q11.2) in three individuals, and del(15)(q11q13) and del(22)(q12.3q13.1) each in two individuals.

**Figure 6.**
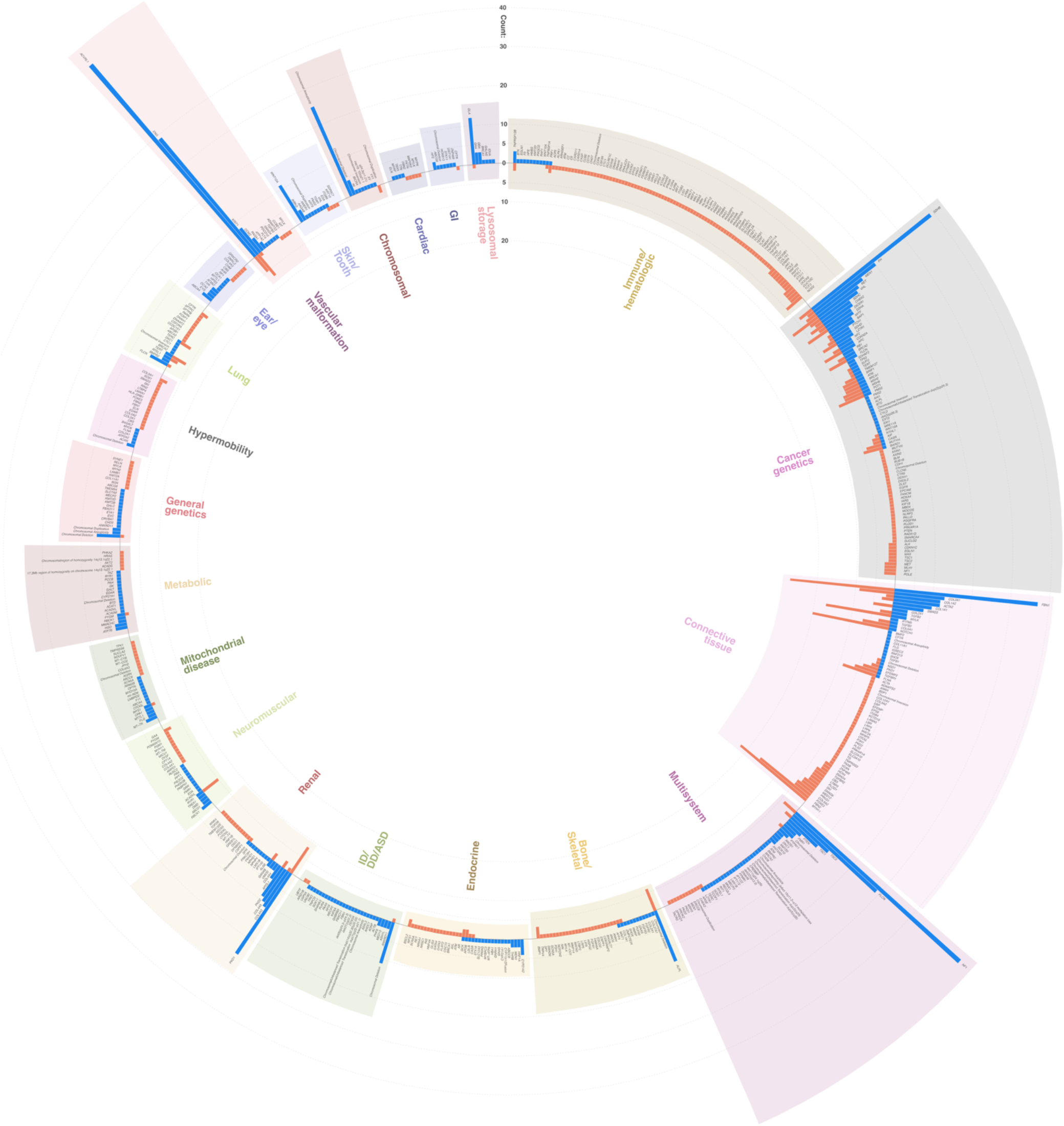
Distribution of Variants Identified by Gene and Clinical Indication Group. Circular bar plot showing how often genetic testing identified a pathogenic variant (blue bars) or a variant of uncertain significance (VUS; orange bars) in each gene, grouped and color-coded by clinical indication group. For each gene, the blue bar height above the baseline represents the total number of pathogenic variants reported, while the orange bar height extending below the baseline represents the total number of VUSs. Each sector is shaded and labeled to show the clinical indication for the visit during which testing was ordered. Reference rings mark variant count intervals for both pathogenic and VUS counts, and gene names appear adjacent to their respective bars.

### WES/WGS patients differ demographically from those receiving other tests

Altogether 168 individuals referred to our clinic during the study period ultimately underwent WES or WGS testing as part of their evaluation; these broad genetic testing modalities made up only 3.96% of all testing ordered and are often inaccessible to adult patients due to insurance denials.^32^ Thus, we sought to identify differences in patient demographics between those patients undergoing WES/WGS (n=168) and those undergoing any other type of testing (n=4,077). Significant differences were observed in clinical indication for referral between the two groups (p=5.51e-98; χ² = 522.5, **Figure S4A-B**). By residual analysis, this difference was primarily driven by an overrepresentation of individuals referred for ID/DD/ASD, general genetics evaluation, and mitochondrial disease among the WES/WGS group. No significant differences were observed in EHR-reported race or sex between the WES/WGS group and other testing group (**Figure S4C-F**). There was a significant difference seen in insurance classes between the two groups (p=1.56e-07; χ² = 34.5), with Medicaid insurance being overrepresented in the WES/WGS group by residual analysis (**Figure S4G-H**). Lastly, patients undergoing WES/WGS were significantly younger (median age 31.0 years) than those who underwent any other genetic testing (median age 43.6 years, p=3.35e-21 by Wilcoxon rank-sum test, **Figure S4I**).

### High diagnostic yield of WES/WGS across all adult patient age groups

WES/WGS tests had a very high diagnostic yield overall (**Figure 7A**). Of the 168 WES/WGS tests completed in the study period, 39.3% returned with a diagnostic pathogenic finding. Non-diagnostic findings were identified in 19.6% of individuals undergoing WES/WGS, including variants of uncertain significance (10.7%), variants indicating unaffected carrier status (7.7%), and variants in candidate genes (1.2%). Altogether, only 41.1% of all WES/WGS tests were completely negative. The diagnostic yield of WES/WGS did not differ based on EHR-reported sex, age, or race (**Figure 7B-D**). Specifically, we observed no significant differences in the odds of identifying pathogenic (p= 0.85; OR 1.00 [0.96–1.03]) or VUS (p=0.80; OR 1.01 [0.95–1.06]) results with increasing patient age, with diagnostic yield above 40% even in patients greater than 55 years old. Comparing the yield of WES/WGS by referral indication (**Figure 7E**), we observed a significantly increased likelihood of obtaining pathogenic results in patients undergoing WES/WGS for ID/DD/ASD compared to all other referral indications (p= 1.46e-04; OR 7.07 [2.70–20.79] by logistic regression).

**Figure 7.**
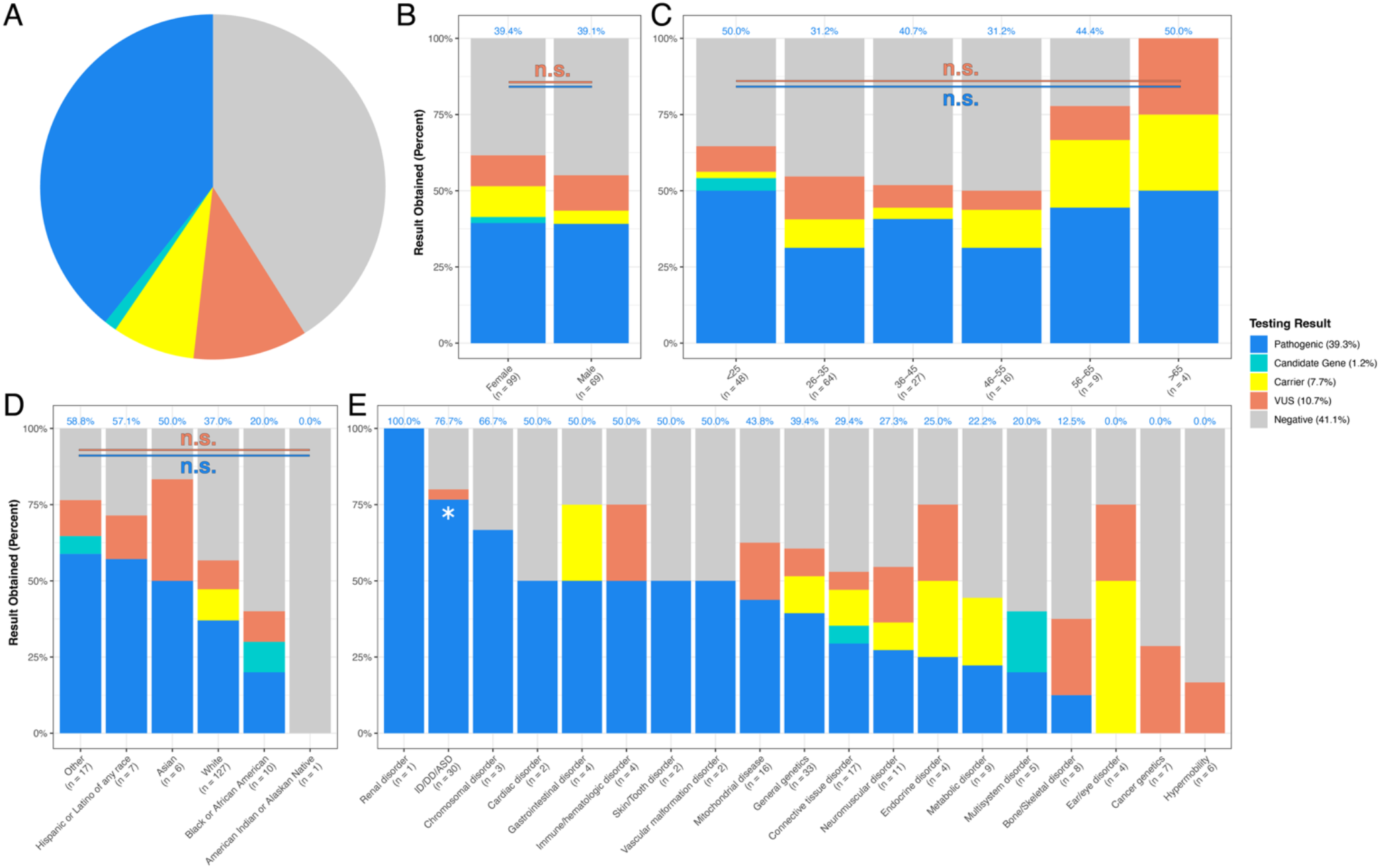
WES/WGS Results by Patient and Visit Characteristics. Results of all 168 WES/WGS tests sent over the study period, stratified by patient and visit characteristics. In all panels, bars indicate the percentage of pathogenic (blue), variant of uncertain significance (VUS; orange), carrier (yellow), candidate gene (aqua), or negative (gray) results. Percent labels in blue above bars show the percent of patients who received a pathogenic result for each subgroup. Sample sizes for each subgroup are shown below the horizontal axis labels for panels B-E. (A) Pie chart showing the overall distribution of results for all 168 patients who underwent WES/WGS during the study period. Percentages for each result classification are shown in the legend. (B) Percent of patients receiving each WES/WGS result classification, stratified by EHR-reported sex. No significant associations were found between sex and the odds of receiving a pathogenic (in blue) or VUS (in orange) result from logistic regression adjusted for age, race, insurance class, and clinical indication group. (C) Percent of patients receiving each WES/WGS result classification, stratified by patient age at visit (plotted in discrete age ranges). No significant associations were found between age (modeled as a continuous variable) and the odds of receiving a pathogenic (in blue) or VUS (in orange) result from logistic regression adjusted for insurance class and clinical indication group. (D) Percent of patients receiving each WES/WGS result classification, stratified by EHR-reported race. No significant differences were found for any racial group, relative to patients with EHR-reported White race, for the odds of receiving either a pathogenic (in blue) or VUS (in orange) result by logistic regression adjusted for age, insurance class, and clinical indication group, accounting for multiple testing across all 6 groups. (E) Percent of patients receiving each WES/WGS result classification, stratified by clinical indication group, ordered by likelihood of receiving a pathogenic result. Indications where WES/WGS testing had a significantly higher odds of identifying a pathogenic result compared to the overall WES/WGS average (ID/DD/ASD) are marked with an asterisk. No significant differences were found for any indication group for the odds of receiving a VUS result. Significance was determined by logistic regression adjusted for age, sex, race, and insurance class, accounting for multiple testing across all 21 groups.

## Discussion

Across eight years and 9,867 visits, our study provides a comprehensive assessment of the scope, diagnostic value, and delivery of care at a single large volume adult general genetics practice, showing how patient characteristics, clinical indications, testing practices, and health-system factors shape this clinical domain. Diagnostic yield remained high throughout adulthood, while patient demographics, testing rates, diagnostic yield, and VUS rate varied substantially by indication. Laboratory utilization shifted alongside changes in cost, coverage, and ordering infrastructure. Diverse care models, including genetic counselor-only visits and expanded telehealth utilization, helped extend the clinic’s reach. Our findings provide some of the first broad practical evidence to inform adult genetics referral pathways, provide guidance for genetic test selection and utilization, and promote diverse models of genetic care for adult patients.

Our work highlights the value of genetic diagnosis well beyond childhood. Pathogenic findings were identified in 24.3% of genetic tests, and diagnostic yield exceeded 17% in every adult age stratum, including patients older than 75 years. Yield was even higher for WES/WGS, which provided diagnoses in 39.3% of adults tested, within the range reported in pediatric WES/WGS sequencing cohorts.^33^ These findings support the implementation of broader access to genetic testing throughout adulthood, with an important caveat: the diagnostic yields we observed are those of testing ordered after evaluation by a genetics-trained clinician, who judged testing to be warranted in only about half of all new patients. Such clinical selection is a central purpose of genetics evaluation, and our diagnostic yield is therefore higher than what would be expected in an unselected population. Additionally, confirmatory and familial-variant testing, which made up >10% of testing sent in our data, which by their nature carry a high pretest expectation of a positive result, likely also contribute to the high diagnostic rates we observe. We also could not systematically distinguish patients who entered care with a molecular diagnosis established elsewhere. Our reported testing yield therefore does not capture the full contribution of genetic diagnosis to the clinic population, which includes both patients undergoing diagnostic evaluation and those receiving care for previously diagnosed conditions. Thus, the high yield we report reflects both the often-underappreciated value of genetic testing in adult patients and the importance of expert-guided patient and test selection. Consequently, our findings should not necessarily be extrapolated to support universal, population-level genetic testing of all adults; rather, expanding access to genetics evaluation and testing requires determining which providers should perform patient and test selection and where it belongs in the care pathway. For some patient groups, the clinical question is already well defined before genetics referral, and testing could be ordered within the referring specialty provider, through a genetic counselor-led visit, or through a health system-level population-based screening or testing program; for other patient groups the diagnostic differential is broad and the choice of test depends on a detailed genetics evaluation. Separating these two groups would allow patients with a well-defined, high-yield presentation to be tested at their point of service without waiting for a clinical genetics appointment, and reserve geneticist evaluation for patients in whom the differential is broad and expert examination and test selection are needed.

Our indication-specific testing rates and yields offer a starting point for developing patient and test selection pathways. Renal and vascular malformation referrals represented high-yield clinical populations: testing was ordered at 69.1% and 57.2% of new-patient visits, respectively, with pathogenic-result rates of 46.1% and 49.2%. More than 98% of testing in both groups consisted of multi-gene panels, single-gene tests, or familial-variant tests – testing methodologies with relatively straightforward ordering and pretest counseling. These same testing modalities predominated in certain lower-yield clinical indication groups, accounting for 96.8% of testing for connective tissue disorders and 95.8% for immune/hematologic disorders, despite pathogenic-result rates of 15.3% and 9.7%, respectively. Similarly, in cancer genetics, where clear testing guidelines exist, testing was sent for 64.0% of new patient visits, more than 98% of which consisted of multigene panels, single gene testing, or familial variant testing, with a 16.3% diagnostic rate. However, these lower yields do not necessarily mean that the testing approach for these clinical indications is poorly defined, nor should they justify withholding clinically indicated testing: even a diagnostic yield of 9.7% represents a substantial opportunity to identify diagnoses that may change patient management. These clinical indications may instead represent an

opportunity for embedding genetic counselors in the specialty clinics that see these patients or incorporating testing into specialty workflows with genetics support to allow testing to proceed where patients already receive care, without requiring every patient to first undergo evaluation in a genetics clinic. These pathways would continue to depend on genetics expertise; the high VUS rates we observed in these low-diagnostic-yield groups – 23.0% for connective tissue disease and 38.1% for immune/hematologic disorders – reinforce the importance of pretest counseling, clinical correlation, and expert support for result interpretation. Thus, both higher- and lower-yield indications with relatively standardized testing approaches present opportunities to broaden the reach of an already-strained genetics workforce, allowing genetics providers to focus on patients who require a detailed genetics examination, expert interpretation of complex findings, or counseling and management following a positive result.

Our data also highlight another central question in adult genetics care: which test to order. NGS gene panels accounted for approximately 63% of tests sent from our clinic, with a diagnostic yield of 16.2%, whereas WES/WGS accounted for approximately 4%, with a significantly higher diagnostic yield of 39.3%. However, the two groups are not directly comparable. The WES/WGS cohort was younger and enriched for ID/DD/ASD, general genetics evaluation, and mitochondrial disease – indications for which a broad test is often the more appropriate choice; thus, the difference in diagnostic yield that we observe does not imply that WES/WGS would outperform gene panel testing across all adult patients. The two modalities also differ in how readily they are ordered. Panels are easier to order, more consistently covered by insurance, and simpler to counsel for, which lowers the threshold for sending them and, in turn, their expected yield; WES/WGS is typically sent only when suspicion of an underlying genetic condition is already very high. These properties cut in different directions for different indications. For many of the lower-yield indication groups in our data, panels are likely the right test, as discussed above, and their more direct counseling and interpretation make them the easier modality to deploy outside the genetics clinic. For other indications, such as ID/DD/ASD, multisystem disorders, and suspected mitochondrial disease, our data support front-line WES/WGS in adults with these presentations, as already embedded in pediatric guidelines.^8^ Adult genetic test-selection guidelines that define when a panel is sufficient and when WES/WGS should be first-line would make these distinctions actionable for clinicians outside genetics, and would help clinicians avoid mistakes in test selection.

Beyond referral pathways and test selection, our data also inform the delivery of adult genetics care. Physician-led visits accounted for approximately three quarters of encounters and nearly all exome, genome, microarray, karyotype, and metabolic testing, yet testing was ordered at only 48.7% of new-patient visits. These patterns highlight the physician geneticist’s role in evaluating whether a presentation is suggestive of genetic disease, using examination to refine the differential, and determining whether and what testing would be informative. Genetic counselor-only visits showed a different pattern: testing was ordered at 80.9% of new-patient visits, more than one quarter of testing ordered was for a known familial variant, and broad or specialized testing was only rarely ordered. This pattern is consistent with directing patients with well-defined testing questions to counselor-led care, where the principal needs are pretest counseling, focused test selection, and result interpretation. Telehealth already supports this approach: approximately half of counselor-only new-patient visits were virtual, and counselor-only visits accounted for nearly three quarters of all new-patient telehealth encounters. Thus, a substantial component of initial genetics care is already being delivered without requiring patients to travel to a tertiary center. Nurse practitioner visits served a complementary role, predominantly providing follow-up for established patients and supporting the ongoing care that genetic conditions require after diagnosis. Together, these patterns offer a practical basis for deliberate triage: physician-led evaluation for complex or unresolved diagnostic questions, counselor-led care for patients with defined testing needs that do not require a physician examination, and advanced practice provider care for ongoing surveillance and management. Expanding these established pathways, with clear triage criteria and access to physician genetics expertise when needed, could broaden the clinic’s reach while preserving physician capacity for complex evaluation and medical management. Realizing these possibilities, however, will require significant policy change: direct billing and consistent reimbursement for genetic counselors, and licensure reforms that allow geneticists and genetic counselors to provide telehealth across state lines.^34,35^

Beyond how care is delivered, our data also speaks to the question of whether genetic evaluation is still worthwhile in older adults. The answer depends on what is ordered. Older adults were significantly more likely to undergo testing after a new-patient evaluation, even as the overall likelihood of a pathogenic result declined with age, from 37% in patients aged 18–25 to 18% in those over 65. Yield nonetheless remained substantial at every age: more than one in six tests returned a pathogenic finding in every adult age stratum. Broad sequencing showed a different pattern entirely. WES/WGS yield did not change with age and remained above 30% in every age group, exceeding 40% in patients over 55. Taken together, these findings suggest that the age-related decline in overall yield reflects a shift in presentation and test selection rather than in the underlying biology: older adults more often present with single-organ-system disease and are more often tested with panels, whereas the complex presentations that prompt broad sequencing retain their yield regardless of age. Although few older adults underwent WES/WGS, and the estimates for that group are correspondingly imprecise, the clinical principle is clear: age alone should neither discourage genetic evaluation nor lead to narrower testing when the presentation warrants a broader approach. The value of genetic evaluation in older adults also extends to younger generations. Testing older relatives for variants identified in younger family members, together with assessment of their clinical features, can help clarify pathogenicity by showing whether those variants segregate with disease within the family;^36^ similarly, identifying a pathogenic variant in an older adult can also guide cascade testing, surveillance, and anticipatory guidance for younger relatives.^16^ Together, these diagnostic and family benefits support genetic evaluation throughout adulthood, with test selection guided by clinical presentation and potential benefit rather than age alone.

Laboratory utilization, by contrast, shifted alongside changes in patient costs, insurance coverage, reflex-testing policies, and EHR ordering infrastructure. Over time, testing became concentrated among laboratories offering affordable testing and streamlined ordering. These findings have major implications:

EHR integration, in-network status, and capped-cost programs influence genetic testing selection, and can be used pro-actively to shape institutional testing patterns. At the same time, these findings suggest that patterns of test and laboratory use reflect the economic reality of the clinic as much as its clinical judgment and should be read with that caveat in mind.

There are several broad limitations to our study that shape how our findings should be interpreted. First, we present a single-center study. Although our practice is among the largest adult general genetics clinics in the country, its referral population, payer mix, and laboratory relationships are specific to one academic health system in one region. Thus, the patterns we describe may not generalize to community practices or to other health systems. Second, adult genetics care is fragmented even within our institution. Routine hereditary cancer, neurogenetics, and cardiac genetics evaluations are largely performed in separate specialty services, so these indications are under-represented in our data relative to their true prevalence, and our findings describe a general adult genetics practice rather than the full scope of genetic testing in adults within our health system. Third, our data capture testing that was sent, not testing that was recommended. Tests that were discussed but declined, denied by insurance, or never completed are not represented, so our testing rates reflect what was ordered and completed rather than what was clinically indicated, and the reasons testing was not pursued could not be characterized. Relatedly, we did not systematically identify new patients who arrived with prior genetic testing already completed; because such patients often do not require additional testing, their presence lowers the apparent proportion of new patients for whom testing was ordered, and the true rate of indicated testing is likely higher than the 52% we report. Finally, we did not measure management changes, patient-reported outcomes, or comparative access and outcomes across care models, so the pathways we propose in this discussion, while based on a broad body of data, will require prospective evaluation.

Taken together, these findings provide a practice-level evidence base for adult genetics that has not previously existed. Although guidelines exist for genetic testing in cancer genetics and selected other specialties, guidance across the broader scope of adult genetics remains limited, often relying on extrapolation from pediatric experience^2,15^ and descriptive studies of adult genetics clinics that have characterized referral volume and indication mix without linking them to testing decisions and outcomes.^21,22^ Our study addresses this gap directly, with data on diagnostic yield across the adult lifespan and across testing modalities; on indication-specific testing rates, test selection, and outcomes; on the division of labor among physician geneticists, genetic counselors, and advanced practice providers; and on the response of referral patterns and laboratory utilization to clinic policy and ordering infrastructure. Adult-specific guidance built on such data will need to be matched by coverage policies that support appropriate genomic testing in adults, by recognition and reimbursement of genetic counselors as independent providers, by genetics education in medical school and in internal and family medicine training, and by implementation infrastructure at the health-system level that is designed with equity and accessibility in mind.^37^ What unites these findings is that the limiting factor in adult genetics is not the value of genetic testing, which was high across indications, modalities, and ages, but the capacity to deliver it. Genetic diagnosis remains relevant throughout the adult lifespan; the task now is to make appropriate evaluation, testing, and follow-up consistently accessible to all the adult patients who stand to benefit.

## Data Availability

All figures, supplementary figures, summary statistics, and the complete R code used to generate them are publicly available as an interactive analysis notebook at https://drivas-lab.github.io/medgenoutcomes-2025/, with source code in the accompanying repository (https://github.com/drivas-lab/medgenoutcomes-2025). Individual-level data, aside from what is included in the manuscript and supplemental materials, cannot be shared because of patient privacy concerns; summary data beyond those presented are available from the corresponding author upon reasonable request.

## Acknowledgments

This study itself did not receive external funding. T.G.D. is supported by the Burroughs Wellcome Fund. K.L.N is supported in part by the Breast Cancer Research Foundation, Basser Center for BRCA, the Gray Foundation, and the National Institutes of Health - R01 CA176785 and R01 CA192393.

## Author Contributions

J.I.G. and T.G.D. conceived and planned the study and analysis plan. J.I.G., C.K., E.T. and T.G.D. carried out the analyses. J.I.G., Y.E., S.A., A.R., C.C., Z.B., I.E., L.H., E.K., T.C., S.C, B.N.G, S.M.G., A.S., K.L.N., M.R., S.K. and T.G.D. contributed to data preparation and analysis. J.I.G., K.L.N., M.R., S.K., and T.G.D. contributed to the interpretation of the results. T.G.D. supervised the project. J.I.G. and T.G.D. took the lead in writing the manuscript. All authors provided critical feedback and helped shape the research, analysis and manuscript.

---

**Figure S1.**
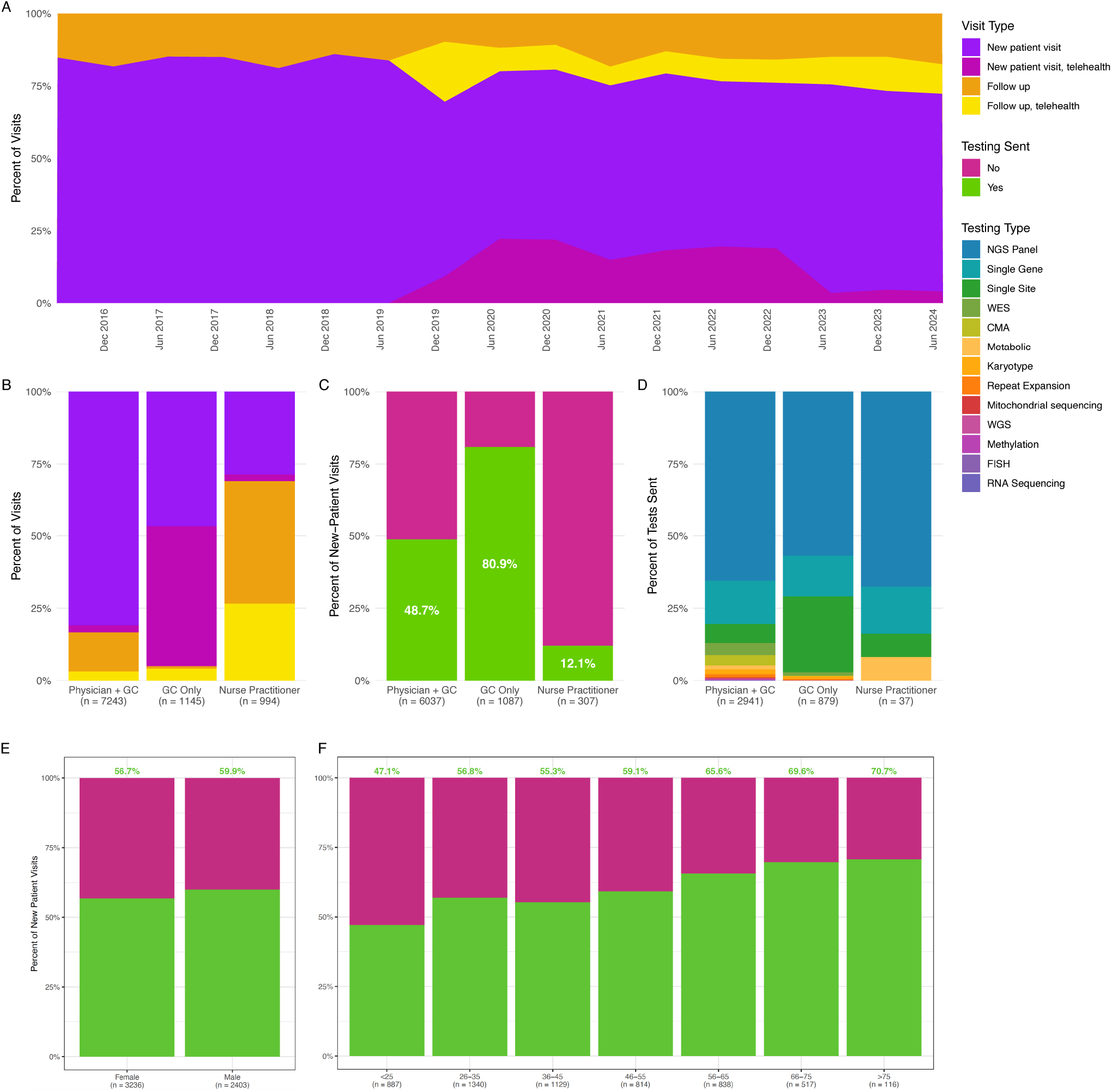
Visit types over time and by provider type. (A) Type of visit (new patient vs. follow-up, in person vs. telehealth) for all visits, shown over time in 6-month intervals from October 1, 2016, to September 30, 2024. Telehealth was introduced in 2020 with the COVID-19 pandemic and associated stay-at-home orders; it has since declined, particularly for new-patient evaluations, but remains a substantial component of follow-up care. (B) Type of visit, by provider type, for all 9,382 visits with a documented provider. Provider types are physician with genetic counselor (n = 7,243), genetic counselor only (n = 1,145), and nurse practitioner (n = 994). Visit types are color-coded as in panel A. (C) Proportion of new-patient visits at which genetic testing was sent, by provider type; the percent tested is shown within each bar. (D) Type of testing sent at new-patient visits, by provider type. (E) Percent of new patient visits during which testing was sent, stratified by EHR-reported sex, with all visits for hypermobility excluded from analysis. Males are significantly more likely to have testing sent; p=0.0169, OR=1.14(1.02-1.26). (F) Percent of new patient visits during which testing was sent, stratified by patient age (plotted in discrete age ranges), with all visits for hypermobility excluded from analysis. Older patients are significantly more likely to have testing sent; p=2.43e-13; OR=1.01(1.01-1.02).

**Figure S2.**
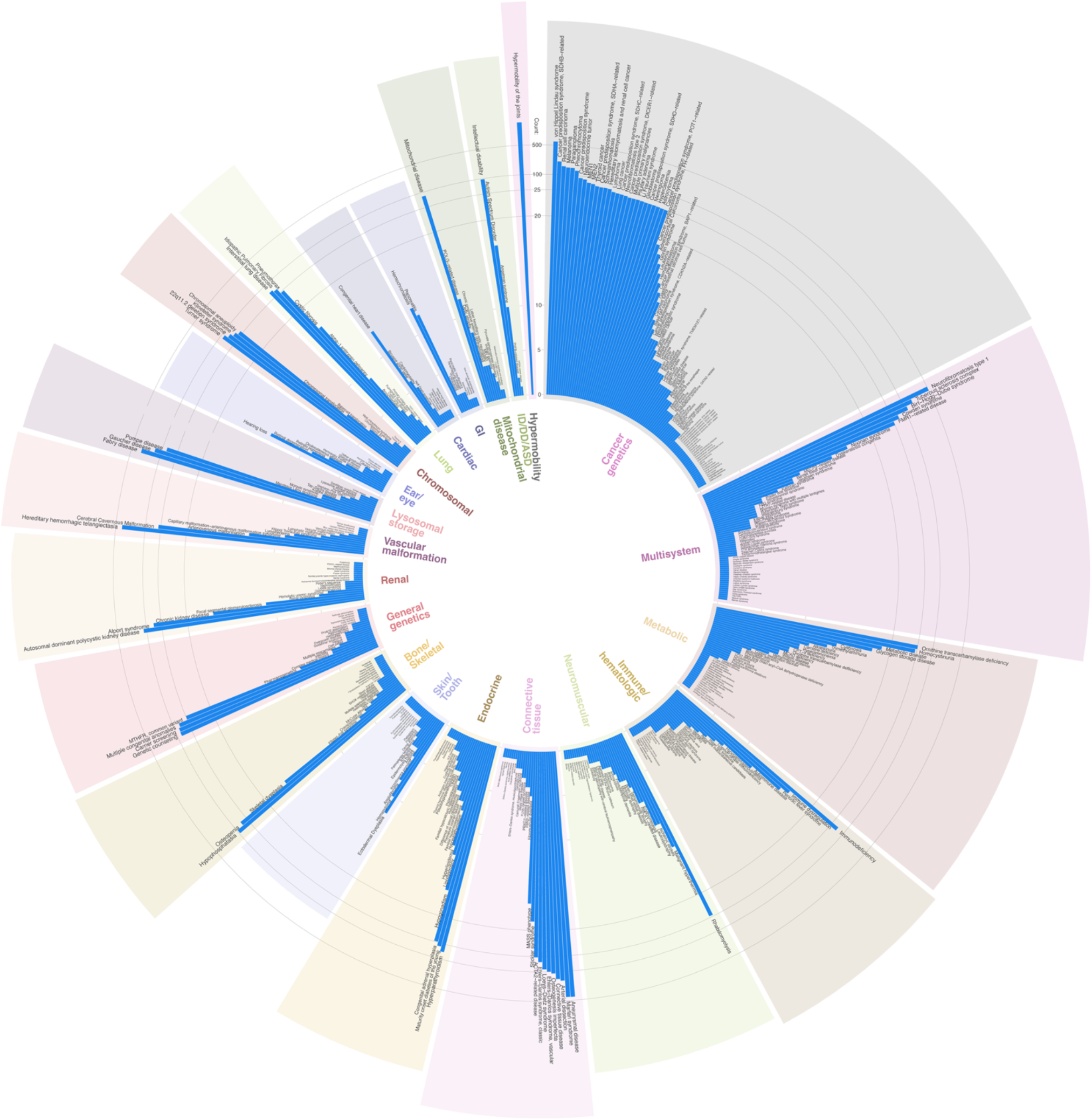
Distribution of Patient Visits by Clinical Indication and Indication Group. Circular bar plot displaying the number of patient visits for each unique clinical indication, grouped and color-coded by indication group, across the study period. Indications are arranged around the circle by indication group, with each indication uniquely assigned to one group. Each blue bar represents a specific indication (e.g., “Marfan syndrome”), and bar height reflects the number of unique patient visits for that indication. For counts greater than 20, bar heights are transformed using the formula: height = 20 + (n)^0.333, where n is the visit count, to improve visualization. Reference rings denote visit counts, with breaks at 5, 10, 20, 25, 100, and 500. Indication names are shown adjacent to their respective bars.

**Figure S3.**
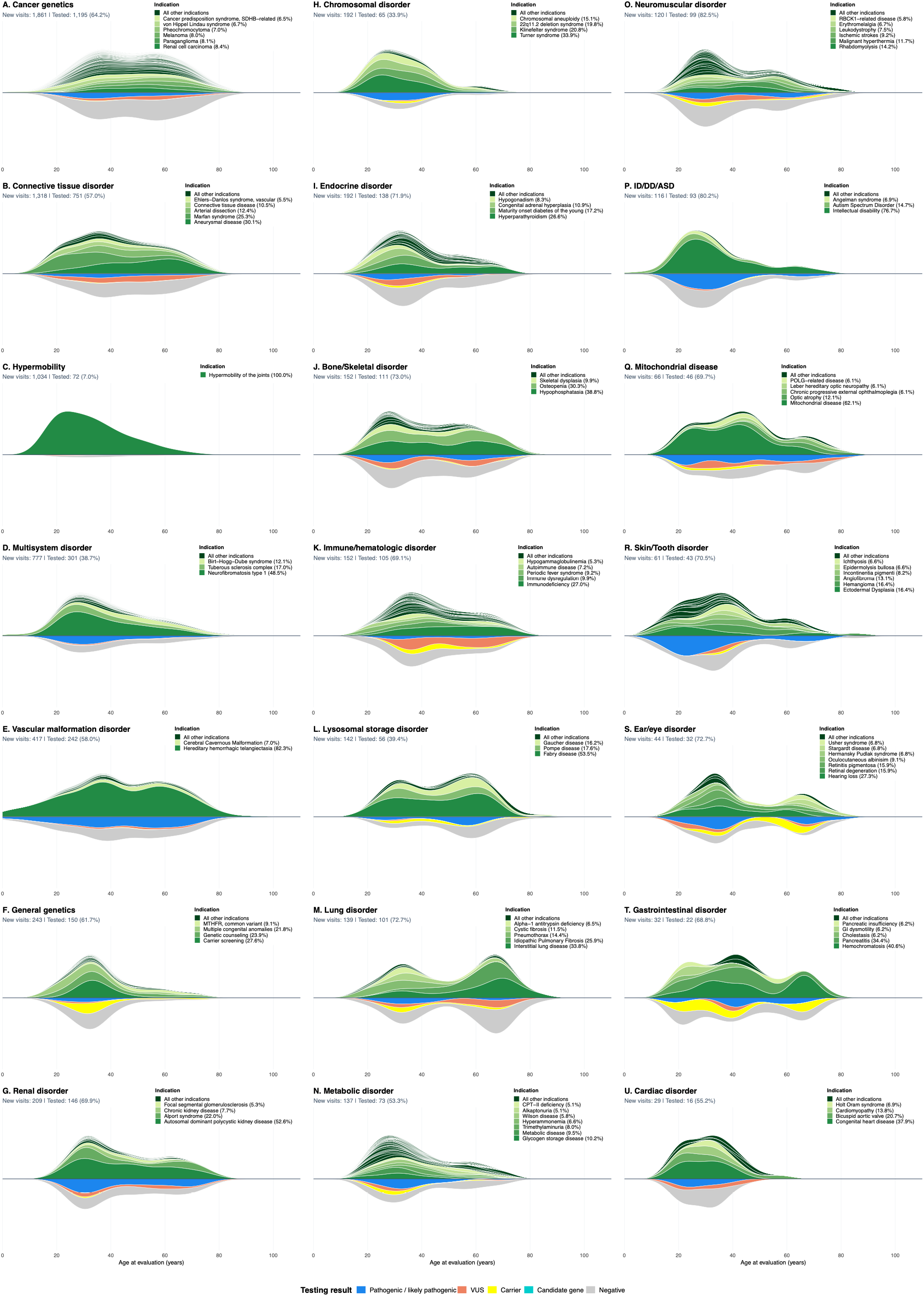
Age distributions of new-patient visits and genetic testing results by indication group. Panels A-U show all 21 indication groups, ordered by decreasing number of new-patient visits. For each group, stacked kernel-density estimates above the horizontal baseline show the distribution of age at evaluation among new-patient visits, including in-person and telehealth encounters. Bands are colored by specific indication within each indication group. Indications representing >5% of visits within an indication group are individually labeled and shown in graded greens; all remaining indications are retained as separate, unlabeled dark-green bands and represented collectively in the legend as “All other indications.” Mirrored stacked densities below the baseline show the age distribution of the subset of new-patient visits for which testing was sent. These bands are colored according to detailed testing classification: pathogenic/likely pathogenic (blue), variant of uncertain significance (VUS; salmon), carrier (yellow), candidate gene (cyan), or negative (gray). Within each panel, the upper density is normalized to have a total area of 1, whereas the lower density has an area equal to the proportion of new-patient visits that underwent testing. Panel subtitles report the number of new visits, the number tested, and the percentage tested. The age axis is shared across panels, while vertical scales vary by indication group.

**Figure S4.**
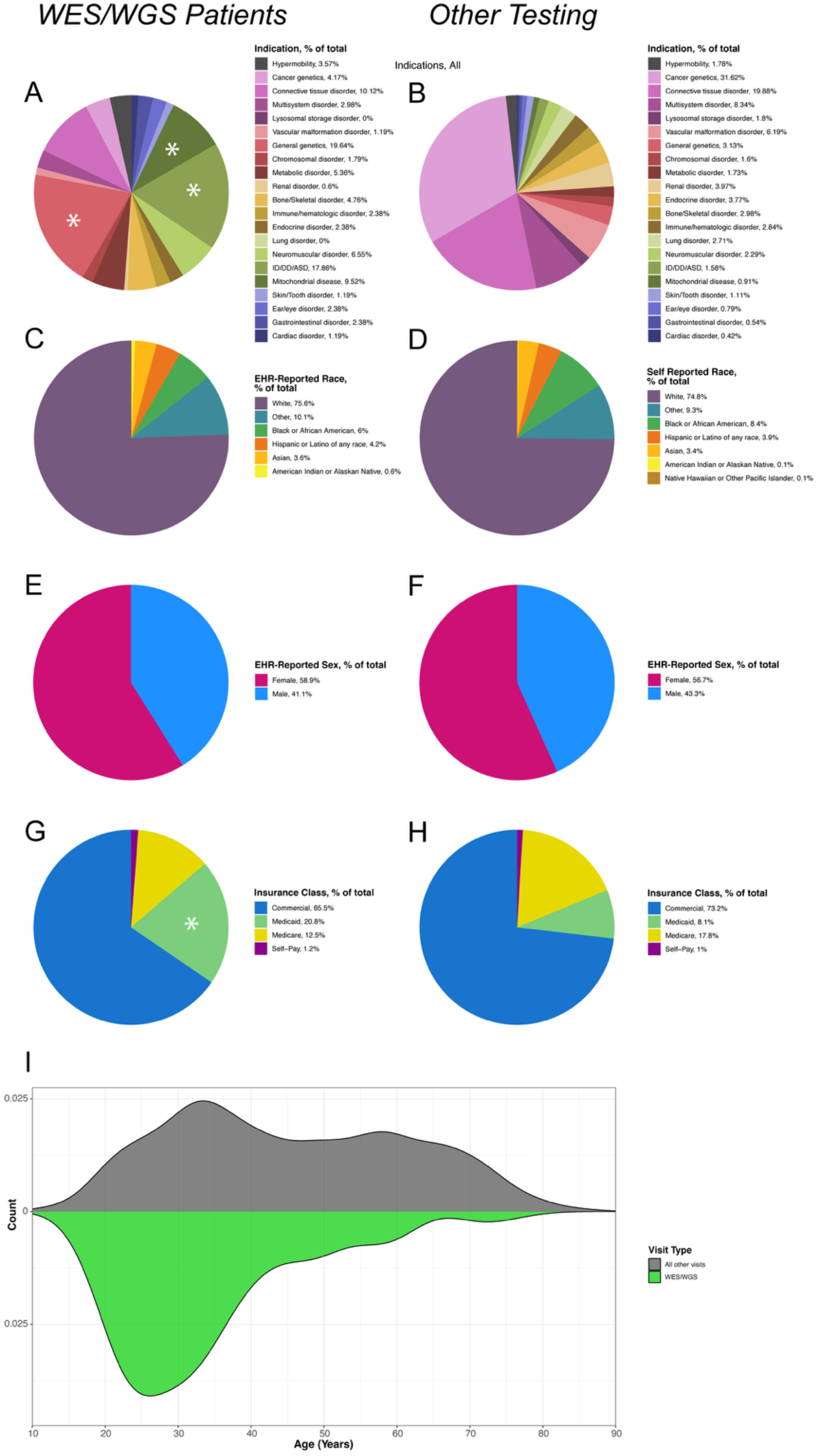
Characteristics of Patients Who Received WES/WGS vs. Other Genetic Tests. Each panel shows the distribution of patient characteristics stratified by whether exome/genome sequencing (WES/WGS, n = 168) was sent (panels A, C, E, G, bottom half of I) or another type of genetic test was sent (n = 4,077, panels B, D, F, H, top half of I). Percentages for each subgroup are indicated in the adjacent legends. (A-B) Proportion of patients by clinical indication group for the visit. (C-D) Proportion of patients by EHR-reported race. (E-F) Proportion of patients by EHR-reported sex. (G-H) Proportion of patients by financial class (insurance type) at the time of the visit. (I) Distribution of patient ages at time of visit. Chi-square analysis comparing WES/WGS to other genetic tests was performed for each patient characteristic in panels A-H. Significant differences were observed for clinical indication (panels A–B) and financial class (panels G–H) only. In those panels, asterisks in the WES/WGS figures indicate categories that were significantly overrepresented among WES/WGS patients, based on residual analysis. Differences in patient age distribution were assessed by Wilcoxon rank-sum test in panel I, where a significant difference was seen with patient ages skewing younger in the WES/WGS group (p=3.35e-21).

**Table S1.** Indication to Indication Group Mappings, with Visit Counts.

| Indication | Indication Group | Count |
| --- | --- | --- |
| Hypophosphatasia | Bone/Skeletal disorder | 93 |
| Osteopenia | Bone/Skeletal disorder | 51 |
| Skeletal dysplasia | Bone/Skeletal disorder | 17 |
| Klippel-Feil syndrome | Bone/Skeletal disorder | 6 |
| Craniosynostosis | Bone/Skeletal disorder | 5 |
| McCune Albright | Bone/Skeletal disorder | 4 |
| Tarlov cyst | Bone/Skeletal disorder | 3 |
| Heterotopic ossification | Bone/Skeletal disorder | 2 |
| Multiple epiphyseal dysplasia | Bone/Skeletal disorder | 2 |
| Osteomalacia | Bone/Skeletal disorder | 2 |
| SHOX-related disease | Bone/Skeletal disorder | 2 |
| Short stature | Bone/Skeletal disorder | 2 |
| Avascular necrosis | Bone/Skeletal disorder | 1 |
| Clubfeet | Bone/Skeletal disorder | 1 |
| Fibrodysplasia Ossificans Progressiva | Bone/Skeletal disorder | 1 |
| Increased bone density | Bone/Skeletal disorder | 1 |
| Mandibuloacral dysplasia | Bone/Skeletal disorder | 1 |
| Osteopathia Striata | Bone/Skeletal disorder | 1 |
| Poland syndrome | Bone/Skeletal disorder | 1 |
| Ribbing disease | Bone/Skeletal disorder | 1 |
| Scoliosis | Bone/Skeletal disorder | 1 |
| Syndactyly | Bone/Skeletal disorder | 1 |
| von Hippel Lindau syndrome | Cancer genetics | 576 |
| Cancer predisposition syndrome, SDHB-related | Cancer genetics | 228 |
| Renal cell carcinoma | Cancer genetics | 175 |
| Melanoma | Cancer genetics | 165 |
| Paraganglioma | Cancer genetics | 163 |
| Pheochromocytoma | Cancer genetics | 140 |
| Cancer predisposition syndrome | Cancer genetics | 86 |
| Neuroendocrine tumor | Cancer genetics | 80 |
| MEN1 | Cancer genetics | 64 |
| MEN2 | Cancer genetics | 60 |
| Thyroid cancer | Cancer genetics | 52 |
| Cancer predisposition syndrome, SDHA-related | Cancer genetics | 50 |
| Schwannomatosis | Cancer genetics | 50 |
| Hereditary leiomyomatosis and renal cell cancer | Cancer genetics | 49 |
| Lymphoma | Cancer genetics | 39 |
| Lung cancer | Cancer genetics | 37 |
| Cancer predisposition syndrome, SDHC-related | Cancer genetics | 36 |
| Neurofibromatosis type 2 | Cancer genetics | 34 |
| Cancer predisposition syndrome, DICER1-related | Cancer genetics | 31 |
| Multiple primary malignancies | Cancer genetics | 31 |
| Pituitary adenoma | Cancer genetics | 31 |
| Li Fraumeni syndrome | Cancer genetics | 30 |
| Glioblastoma | Cancer genetics | 29 |
| Cancer predisposition syndrome, SDHD-related | Cancer genetics | 28 |
| Meningioma | Cancer genetics | 28 |
| Hypercalcemia | Cancer genetics | 27 |
| Astrocytoma | Cancer genetics | 26 |
| Cancer predisposition syndrome, POT1-related | Cancer genetics | 25 |
| Cancer predisposition syndrome, FH-related | Cancer genetics | 19 |
| Adrenocortical Carcinoma | Cancer genetics | 18 |
| Gorlin syndrome | Cancer genetics | 17 |
| Leukemia | Cancer genetics | 16 |
| Hemangioblastoma | Cancer genetics | 15 |
| Cancer predisposition syndrome, BAP1-related | Cancer genetics | 14 |
| Gastrointestinal stromal cell tumor | Cancer genetics | 13 |
| Carcinoid tumor | Cancer genetics | 11 |
| Basal cell carcinoma | Cancer genetics | 10 |
| Cancer predisposition syndrome, CDKN2A-related | Cancer genetics | 9 |
| Mesothelioma | Cancer genetics | 9 |
| Myelodysplastic syndrome | Cancer genetics | 9 |
| Pancreatic cancer | Cancer genetics | 8 |
| Lipomatosis | Cancer genetics | 7 |
| Multiple myeloma | Cancer genetics | 7 |
| Osteochondroma | Cancer genetics | 7 |
| Sarcoma | Cancer genetics | 7 |
| Brain cancer | Cancer genetics | 6 |
| Breast cancer | Cancer genetics | 6 |
| Retinoblastoma | Cancer genetics | 6 |
| Cancer predisposition syndrome, TMEM127-related | Cancer genetics | 5 |
| Carney Complex | Cancer genetics | 5 |
| Gastrinoma | Cancer genetics | 5 |
| RET-related disease | Cancer genetics | 5 |
| Testicular cancer | Cancer genetics | 5 |
| Wilms tumor | Cancer genetics | 5 |
| Adrenal Adenoma | Cancer genetics | 4 |
| Colon cancer | Cancer genetics | 4 |
| Seminoma | Cancer genetics | 4 |
| Adenocarcinoma of the esophagus | Cancer genetics | 3 |
| Ependymoma | Cancer genetics | 3 |
| Lynch syndrome | Cancer genetics | 3 |
| Renal angiomyolipoma | Cancer genetics | 3 |
| Cancer predisposition syndrome, GATA2-related | Cancer genetics | 2 |
| Cylindroma | Cancer genetics | 2 |
| Ganglioneuroma | Cancer genetics | 2 |
| Lymphangiomatosis | Cancer genetics | 2 |
| Osteoma | Cancer genetics | 2 |
| RUNX1-related disease | Cancer genetics | 2 |
| Squamous cell carcinoma | Cancer genetics | 2 |
| Thymoma | Cancer genetics | 2 |
| Angiomyolipoma | Cancer genetics | 1 |
| Cancer predisposition syndrome, ATM-related | Cancer genetics | 1 |
| Cancer predisposition syndrome, CHEK2-related | Cancer genetics | 1 |
| Cancer predisposition syndrome, DDX41-related | Cancer genetics | 1 |
| Cancer predisposition syndrome, FANCA-related | Cancer genetics | 1 |
| Cancer predisposition syndrome, MITF-related | Cancer genetics | 1 |
| Cervical cancer | Cancer genetics | 1 |
| Chondrosarcoma | Cancer genetics | 1 |
| Chordoma | Cancer genetics | 1 |
| Colon polyps | Cancer genetics | 1 |
| Cystadenoma | Cancer genetics | 1 |
| Familial adenomatous polyposis | Cancer genetics | 1 |
| Hemangioendothelioma | Cancer genetics | 1 |
| Hepatocellular carcinoma | Cancer genetics | 1 |
| Inflammatory myofibroblastic tumor | Cancer genetics | 1 |
| Leiomyosarcoma | Cancer genetics | 1 |
| MEN4 | Cancer genetics | 1 |
| Ovarian cancer | Cancer genetics | 1 |
| Prolactinoma | Cancer genetics | 1 |
| Prostate cancer | Cancer genetics | 1 |
| Renal oncocytoma | Cancer genetics | 1 |
| Rhabdomyosarcoma | Cancer genetics | 1 |
| Skin cancer | Cancer genetics | 1 |
| Trichoepitheliomas | Cancer genetics | 1 |
| Urothelial carcinoma | Cancer genetics | 1 |
| Uterine cancer | Cancer genetics | 1 |
| Congenital heart disease | Cardiac disorder | 13 |
| Bicuspid aortic valve | Cardiac disorder | 6 |
| Cardiomyopathy | Cardiac disorder | 5 |
| Holt Oram syndrome | Cardiac disorder | 2 |
| Aortic coarctation | Cardiac disorder | 1 |
| Mitral Valve Prolapse | Cardiac disorder | 1 |
| Patent foramen ovale | Cardiac disorder | 1 |
| Pulmonic valve stenosis | Cardiac disorder | 1 |
| Scimitar Syndrome | Cardiac disorder | 1 |
| Valvular heart disease | Cardiac disorder | 1 |
| Turner syndrome | Chromosomal disorder | 80 |
| 22q11.2 deletion syndrome | Chromosomal disorder | 59 |
| Klinefelter syndrome | Chromosomal disorder | 44 |
| Chromosomal aneuploidy | Chromosomal disorder | 33 |
| Chromosomal translocation | Chromosomal disorder | 8 |
| Balanced translocation | Chromosomal disorder | 5 |
| Trisomy 21 | Chromosomal disorder | 3 |
| Chromosomal inversion | Chromosomal disorder | 2 |
| Wolf-Hirschhorn syndrome | Chromosomal disorder | 2 |
| Trisomy 18 | Chromosomal disorder | 1 |
| Trisomy 8 | Chromosomal disorder | 1 |
| Aneurysmal disease | Connective tissue disorder | 427 |
| Marfan syndrome | Connective tissue disorder | 424 |
| Arterial dissection | Connective tissue disorder | 181 |
| Connective tissue disease | Connective tissue disorder | 162 |
| Osteogenesis imperfecta | Connective tissue disorder | 112 |
| Ehlers-Danlos syndrome, vascular | Connective tissue disorder | 98 |
| Loeys-Dietz syndrome | Connective tissue disorder | 82 |
| Ehlers-Danlos syndrome, classic | Connective tissue disorder | 44 |
| ACTA2-related disease | Connective tissue disorder | 27 |
| Stickler syndrome | Connective tissue disorder | 19 |
| MASS phenotype | Connective tissue disorder | 17 |
| Fibromuscular dysplasia | Connective tissue disorder | 5 |
| FBN2-related disease | Connective tissue disorder | 4 |
| PRKG1-related disease | Connective tissue disorder | 4 |
| FLNA-related syndrome | Connective tissue disorder | 3 |
| Moya-moya disease | Connective tissue disorder | 3 |
| Camurati Engelmann syndrome | Connective tissue disorder | 2 |
| Contractures | Connective tissue disorder | 2 |
| Ehlers-Danlos syndrome, musculocontractural | Connective tissue disorder | 2 |
| Bifid uvula | Connective tissue disorder | 1 |
| Dupuytren contractures | Connective tissue disorder | 1 |
| ELN-related disease | Connective tissue disorder | 1 |
| Ehlers-Danlos syndrome, kyphoscoliotic | Connective tissue disorder | 1 |
| Hernia | Connective tissue disorder | 1 |
| Spinal hygroma | Connective tissue disorder | 1 |
| Weill-Marchesani syndrome | Connective tissue disorder | 1 |
| Hearing loss | Ear/eye disorder | 14 |
| Retinal degeneration | Ear/eye disorder | 9 |
| Retinitis pigmentosa | Ear/eye disorder | 7 |
| Oculocutaneous albinism | Ear/eye disorder | 5 |
| Hermansky Pudlak syndrome | Ear/eye disorder | 3 |
| Stargardt disease | Ear/eye disorder | 3 |
| Usher syndrome | Ear/eye disorder | 3 |
| Cataract | Ear/eye disorder | 2 |
| Axenfield-Reiger syndrome | Ear/eye disorder | 1 |
| Choroideremia | Ear/eye disorder | 1 |
| Occular albinism | Ear/eye disorder | 1 |
| Hyperparathyroidism | Endocrine disorder | 55 |
| Maturity onset diabetes of the young | Endocrine disorder | 35 |
| Congenital adrenal hyperplasia | Endocrine disorder | 23 |
| Hypogonadism | Endocrine disorder | 17 |
| Lipodystrophy | Endocrine disorder | 13 |
| Hyperlipidemia | Endocrine disorder | 12 |
| Hypoparathyroidism | Endocrine disorder | 8 |
| Familial hypercholesterolemia | Endocrine disorder | 7 |
| Premature ovarian failure | Endocrine disorder | 7 |
| Difference of sexual development | Endocrine disorder | 6 |
| Hypokalemia | Endocrine disorder | 6 |
| Familial hypocalciuric hypercalcemia | Endocrine disorder | 5 |
| Hyperaldosteronism | Endocrine disorder | 4 |
| Obesity | Endocrine disorder | 4 |
| Pseudohypoparathyroidism | Endocrine disorder | 4 |
| Thyroid hormone resistance syndrome | Endocrine disorder | 3 |
| Androgen insensitivity syndrome | Endocrine disorder | 2 |
| Cushing syndrome | Endocrine disorder | 2 |
| Hypocalcemia | Endocrine disorder | 2 |
| ACE Hyperactivity | Endocrine disorder | 1 |
| Calcinosis | Endocrine disorder | 1 |
| Endocrine disorder | Endocrine disorder | 1 |
| Hyperinsulinism | Endocrine disorder | 1 |
| Hypobetalipoproteinemia | Endocrine disorder | 1 |
| Pseudohypoaldosteronism | Endocrine disorder | 1 |
| Hemochromatosis | Gastrointestinal disorder | 13 |
| Pancreatitis | Gastrointestinal disorder | 12 |
| Cholestasis | Gastrointestinal disorder | 2 |
| GI dysmotility | Gastrointestinal disorder | 2 |
| Hirschsprung disease | Gastrointestinal disorder | 2 |
| Pancreatic insufficiency | Gastrointestinal disorder | 2 |
| Abdominal Pain | Gastrointestinal disorder | 1 |
| Cirrhosis | Gastrointestinal disorder | 1 |
| Hyperbilirubinemia | Gastrointestinal disorder | 1 |
| Non-alcoholic steatohepatitis | Gastrointestinal disorder | 1 |
| Genetic counseling | General genetics | 72 |
| Carrier screening | General genetics | 70 |
| Multiple congenital anomalies | General genetics | 61 |
| MTHFR, common variant | General genetics | 24 |
| Pharmacogenetics | General genetics | 9 |
| Complex medical history | General genetics | 5 |
| Infertility | General genetics | 5 |
| Multiple miscarriages | General genetics | 4 |
| Cleft palate | General genetics | 3 |
| Fatigue | General genetics | 3 |
| Goldenhar syndrome | General genetics | 3 |
| Overgrowth disorder | General genetics | 3 |
| Hemihypertrophy | General genetics | 2 |
| Hydrocephalus | General genetics | 2 |
| PHACE association | General genetics | 2 |
| Aquagenic palm wrinkling | General genetics | 1 |
| Cyclic vomiting | General genetics | 1 |
| Dysmorphism syndrome | General genetics | 1 |
| Hyperhidrosis | General genetics | 1 |
| Swyer syndrome | General genetics | 1 |
| Testicular abnormalities | General genetics | 1 |
| Intellectual disability | ID/DD/ASD | 94 |
| Autism Spectrum Disorder | ID/DD/ASD | 17 |
| Angelman syndrome | ID/DD/ASD | 10 |
| PHF8-related disease | ID/DD/ASD | 2 |
| KAT6A-related disease | ID/DD/ASD | 1 |
| Immunodeficiency | Immune/hematologic disorder | 53 |
| Immune dysregulation | Immune/hematologic disorder | 17 |
| Periodic fever syndrome | Immune/hematologic disorder | 16 |
| Autoimmune disease | Immune/hematologic disorder | 12 |
| Hypogammaglobulinemia | Immune/hematologic disorder | 8 |
| Common variable immunodeficiency | Immune/hematologic disorder | 7 |
| Hyper IgE syndrome | Immune/hematologic disorder | 6 |
| Recurrent mucocutaneous candidiasis | Immune/hematologic disorder | 5 |
| Hemophilia | Immune/hematologic disorder | 4 |
| G6PD deficiency | Immune/hematologic disorder | 3 |
| Neutropenia | Immune/hematologic disorder | 3 |
| Polycythemia vera | Immune/hematologic disorder | 3 |
| Porphyria | Immune/hematologic disorder | 3 |
| Recurrent infections | Immune/hematologic disorder | 3 |
| Wiskott-Aldrich syndrome | Immune/hematologic disorder | 3 |
| Bleeding diathesis | Immune/hematologic disorder | 2 |
| Chronic granulomatous disease | Immune/hematologic disorder | 2 |
| Hereditary angioedema | Immune/hematologic disorder | 2 |
| Hidradenitis suppurativa | Immune/hematologic disorder | 2 |
| Hyper IgM syndrome | Immune/hematologic disorder | 2 |
| PAPASH syndrome | Immune/hematologic disorder | 2 |
| Thalassemia | Immune/hematologic disorder | 2 |
| Anemia | Immune/hematologic disorder | 1 |
| Beta thalassemia | Immune/hematologic disorder | 1 |
| Blau syndrome | Immune/hematologic disorder | 1 |
| Bruising | Immune/hematologic disorder | 1 |
| Cytopenia | Immune/hematologic disorder | 1 |
| Diamond Blackfan Anemia | Immune/hematologic disorder | 1 |
| Hemophagocytic lymphohistiocytosis | Immune/hematologic disorder | 1 |
| Hyperferritinemia | Immune/hematologic disorder | 1 |
| Mast cell activation syndrome | Immune/hematologic disorder | 1 |
| TRAPS-related disease | Immune/hematologic disorder | 1 |
| Interstitial lung disease | Lung disorder | 50 |
| Idiopathic Pulmonary Fibrosis | Lung disorder | 41 |
| Pneumothorax | Lung disorder | 22 |
| Cystic fibrosis | Lung disorder | 17 |
| Alpha-1 antitrypsin deficiency | Lung disorder | 9 |
| Bronchiectasis | Lung disorder | 4 |
| Cystic lung disease | Lung disorder | 3 |
| Pulmonary hypertension | Lung disorder | 3 |
| Primary ciliary dyskinesia | Lung disorder | 2 |
| TERC-related disease | Lung disorder | 1 |
| Fabry disease | Lysosomal storage disorder | 315 |
| Gaucher disease | Lysosomal storage disorder | 156 |
| Pompe disease | Lysosomal storage disorder | 71 |
| Maroteaux Lamy syndrome | Lysosomal storage disorder | 6 |
| Morquio syndrome | Lysosomal storage disorder | 6 |
| Krabbe disease | Lysosomal storage disorder | 4 |
| Tay-Sachs disease | Lysosomal storage disorder | 4 |
| Niemann-Pick Type C | Lysosomal storage disorder | 3 |
| Hunter syndrome | Lysosomal storage disorder | 2 |
| Lysosomal storage disease | Lysosomal storage disorder | 2 |
| Sanfilippo syndrome | Lysosomal storage disorder | 2 |
| Scheie disease | Lysosomal storage disorder | 1 |
| Ornithine transcarbamylase deficiency | Metabolic disorder | 25 |
| Homocystinuria | Metabolic disorder | 20 |
| Metabolic disease | Metabolic disorder | 19 |
| Glycogen storage disease | Metabolic disorder | 18 |
| Cystinosis | Metabolic disorder | 14 |
| Trimethylaminuria | Metabolic disorder | 13 |
| Alkaptonuria | Metabolic disorder | 11 |
| Carnitine deficiency | Metabolic disorder | 10 |
| Hyperammonemia | Metabolic disorder | 10 |
| CPT-II deficiency | Metabolic disorder | 9 |
| Ornithine transcarbamylase deficiency | Metabolic disorder | 9 |
| MCAD deficiency | Metabolic disorder | 8 |
| Wilson disease | Metabolic disorder | 8 |
| Methylmalonic acidemia | Metabolic disorder | 7 |
| Copper metabolism disorder | Metabolic disorder | 5 |
| Biotinidase deficiency | Metabolic disorder | 4 |
| Phenylketonuria | Metabolic disorder | 4 |
| Very long chain acyl-CoA dehydrogenase deficiency | Metabolic disorder | 4 |
| CPS1 deficiency | Metabolic disorder | 3 |
| Citrullinemia | Metabolic disorder | 3 |
| Glutaric aciduria | Metabolic disorder | 3 |
| Propionic acidemia | Metabolic disorder | 3 |
| Pseudoxanthoma elasticum | Metabolic disorder | 3 |
| Galactosemia | Metabolic disorder | 2 |
| 11-beta hydroxylase deficiency | Metabolic disorder | 1 |
| ACAD9 deficiency | Metabolic disorder | 1 |
| ASA lyase deficiency | Metabolic disorder | 1 |
| B-ketothiolase deficiency | Metabolic disorder | 1 |
| Barth syndrome | Metabolic disorder | 1 |
| Butyrylcholinesterase deficiency | Metabolic disorder | 1 |
| Canavan disease | Metabolic disorder | 1 |
| Cerebrotendinous Xanthomatosis | Metabolic disorder | 1 |
| Cobalamin C deficiency | Metabolic disorder | 1 |
| Congenital Disorder of Glycosylation | Metabolic disorder | 1 |
| Fatty acid oxidation disorder | Metabolic disorder | 1 |
| GM1 gangliosidosis | Metabolic disorder | 1 |
| Glycerol Kinase Deficiency | Metabolic disorder | 1 |
| Hereditary fructose intolerance | Metabolic disorder | 1 |
| Hurler syndrome | Metabolic disorder | 1 |
| Hyperoxaluria | Metabolic disorder | 1 |
| Lesch Nyhan syndrome | Metabolic disorder | 1 |
| Multiple acyl-CoA dehydrogenase deficiency (MADD) | Metabolic disorder | 1 |
| Zellweger spectrum disorder | Metabolic disorder | 1 |
| Mitochondrial disease | Mitochondrial disease | 73 |
| POLG-related disease | Mitochondrial disease | 12 |
| Optic atrophy | Mitochondrial disease | 8 |
| Chronic progressive external ophthalmoplegia | Mitochondrial disease | 4 |
| Leber hereditary optic neuropathy | Mitochondrial disease | 4 |
| MELAS | Mitochondrial disease | 4 |
| Wolfram syndrome | Mitochondrial disease | 3 |
| Pyruvate dehydrogenase deficiency | Mitochondrial disease | 2 |
| C1QBP-related myopathy | Mitochondrial disease | 1 |
| Maternally inherited diabetes and deafness | Mitochondrial disease | 1 |
| Neurofibromatosis type 1 | Multisystem disorder | 499 |
| Tuberous sclerosis complex | Multisystem disorder | 186 |
| Birt-Hogg-Dube syndrome | Multisystem disorder | 120 |
| Cowden syndrome | Multisystem disorder | 48 |
| FMR1-related disease | Multisystem disorder | 25 |
| Noonan syndrome | Multisystem disorder | 17 |
| Dyskeratosis congenita | Multisystem disorder | 15 |
| MECP2-related disease | Multisystem disorder | 11 |
| Bardet Biedl syndrome | Multisystem disorder | 10 |
| Jacobsen syndrome | Multisystem disorder | 10 |
| Kallman syndrome | Multisystem disorder | 8 |
| Prader Willi syndrome | Multisystem disorder | 6 |
| Sensebrenner syndrome | Multisystem disorder | 6 |
| KBG syndrome | Multisystem disorder | 5 |
| LZTR1-related disease | Multisystem disorder | 5 |
| Noonan syndrome with multiple lentigines | Multisystem disorder | 5 |
| Rubenstein Taybi syndrome | Multisystem disorder | 5 |
| Waardenburg syndrome | Multisystem disorder | 5 |
| Kabuki syndrome | Multisystem disorder | 4 |
| Nail-Patella syndrome | Multisystem disorder | 4 |
| Phelan-McDermid Syndrome | Multisystem disorder | 4 |
| Williams syndrome | Multisystem disorder | 4 |
| Branchio-oto-renal syndrome | Multisystem disorder | 2 |
| Cardiofaciocutaneous syndrome | Multisystem disorder | 2 |
| Chondrodysplasia punctata | Multisystem disorder | 2 |
| Coffin-Siris syndrome | Multisystem disorder | 2 |
| Heterotaxy | Multisystem disorder | 2 |
| Joubert syndrome | Multisystem disorder | 2 |
| Potocki Lupski syndrome | Multisystem disorder | 2 |
| Schwachman Diamond syndrome | Multisystem disorder | 2 |
| TTR Amyloidosis | Multisystem disorder | 2 |
| Treacher-Collins syndrome | Multisystem disorder | 2 |
| Trichorhinophalangeal syndrome | Multisystem disorder | 2 |
| ADNP-related disease | Multisystem disorder | 1 |
| Alagile syndrome | Multisystem disorder | 1 |
| Baraitser-Winter syndrome | Multisystem disorder | 1 |
| Beckwith-Wiedemann syndrome | Multisystem disorder | 1 |
| Cockayne syndrome | Multisystem disorder | 1 |
| Currarino syndrome | Multisystem disorder | 1 |
| Darier disease | Multisystem disorder | 1 |
| Fanconi Anemia | Multisystem disorder | 1 |
| Freeman-Sheldon syndrome | Multisystem disorder | 1 |
| Hajdu-Cheney syndrome | Multisystem disorder | 1 |
| Inherited systemic hyalinosis | Multisystem disorder | 1 |
| Kleefstra syndrome | Multisystem disorder | 1 |
| Legius syndrome | Multisystem disorder | 1 |
| Luscan-Lumish syndrome | Multisystem disorder | 1 |
| Opitz G/BBB Syndrome | Multisystem disorder | 1 |
| Rett Syndrome | Multisystem disorder | 1 |
| Rothmund-Thomson syndrome | Multisystem disorder | 1 |
| Sotos syndrome | Multisystem disorder | 1 |
| VACTERL | Multisystem disorder | 1 |
| Weaver syndrome | Multisystem disorder | 1 |
| Werner syndrome | Multisystem disorder | 1 |
| Rhabdomyolysis | Neuromuscular disorder | 21 |
| Malignant hyperthermia | Neuromuscular disorder | 14 |
| Ischemic strokes | Neuromuscular disorder | 11 |
| Leukodystrophy | Neuromuscular disorder | 11 |
| Erythromelalgia | Neuromuscular disorder | 8 |
| RBCK1-related disease | Neuromuscular disorder | 8 |
| Spinomuscular atrophy | Neuromuscular disorder | 6 |
| Brain small vessel disease | Neuromuscular disorder | 5 |
| Muscular dystrophy | Neuromuscular disorder | 5 |
| Myopathy | Neuromuscular disorder | 5 |
| Frontotemporal dementia | Neuromuscular disorder | 4 |
| Arthrogryposis | Neuromuscular disorder | 3 |
| Ataxia | Neuromuscular disorder | 3 |
| Dysautonomia | Neuromuscular disorder | 3 |
| Polymicrogyria | Neuromuscular disorder | 3 |
| Agenesis of the corpus callosum | Neuromuscular disorder | 2 |
| Cerebral palsy | Neuromuscular disorder | 2 |
| Neuromuscular disease | Neuromuscular disorder | 2 |
| Retinal vasculopathy with cerebral | Neuromuscular disorder | 2 |
| Spasticity | Neuromuscular disorder | 2 |
| Catatonia | Neuromuscular disorder | 1 |
| Charcot Marie Tooth | Neuromuscular disorder | 1 |
| Congenital Central Hypoventilation Syndrome | Neuromuscular disorder | 1 |
| Muscle hypertrophy | Neuromuscular disorder | 1 |
| Myotonia congenita | Neuromuscular disorder | 1 |
| Myotonic dystrophy | Neuromuscular disorder | 1 |
| Neuropathy | Neuromuscular disorder | 1 |
| Psychosis | Neuromuscular disorder | 1 |
| ROBO4-related disorder | Neuromuscular disorder | 1 |
| Seizures | Neuromuscular disorder | 1 |
| Stroke | Neuromuscular disorder | 1 |
| Autosomal dominant polycystic kidney disease | Renal disorder | 123 |
| Alport syndrome | Renal disorder | 54 |
| Chronic kidney disease | Renal disorder | 17 |
| Focal segmental glomerulosclerosis | Renal disorder | 11 |
| Renal dysplasia | Renal disorder | 5 |
| Hemolytic uremic syndrome | Renal disorder | 4 |
| Glomerulonephritis | Renal disorder | 2 |
| Liddle syndrome | Renal disorder | 2 |
| Nephrolithiasis | Renal disorder | 2 |
| Potter's sequence | Renal disorder | 2 |
| Autosomal dominant tubulointerstitial kidney disease | Renal disorder | 1 |
| Bartter syndrome | Renal disorder | 1 |
| Familial juvenile hyperuricaemic nephropathy | Renal disorder | 1 |
| Frasier syndrome | Renal disorder | 1 |
| Geller syndrome | Renal disorder | 1 |
| Minimal change disease | Renal disorder | 1 |
| Nephrocalcinosis | Renal disorder | 1 |
| PODXL-related disease | Renal disorder | 1 |
| Proteinuria | Renal disorder | 1 |
| Ectodermal Dysplasia | Skin/Tooth disorder | 12 |
| Hemangioma | Skin/Tooth disorder | 10 |
| Angiofibroma | Skin/Tooth disorder | 8 |
| Incontinentia pigmenti | Skin/Tooth disorder | 5 |
| Angioma | Skin/Tooth disorder | 4 |
| Epidermolysis bullosa | Skin/Tooth disorder | 4 |
| Ichthyosis | Skin/Tooth disorder | 4 |
| Palmoplantar keratoderma | Skin/Tooth disorder | 3 |
| Dental anomalies | Skin/Tooth disorder | 2 |
| Hypodontia | Skin/Tooth disorder | 2 |
| Piebaldism | Skin/Tooth disorder | 2 |
| Alopecia | Skin/Tooth disorder | 1 |
| Anhidrosis | Skin/Tooth disorder | 1 |
| Epidermal Inclusion Cysts | Skin/Tooth disorder | 1 |
| Epidermodysplasia verruciformis | Skin/Tooth disorder | 1 |
| Epidermolytic hyperkeratosis | Skin/Tooth disorder | 1 |
| Erythrodysplasia verruciformis | Skin/Tooth disorder | 1 |
| Keratoderma | Skin/Tooth disorder | 1 |
| Lentigines | Skin/Tooth disorder | 1 |
| Pruritis | Skin/Tooth disorder | 1 |
| Pustular psoriasis | Skin/Tooth disorder | 1 |
| Trichothiodystrophy | Skin/Tooth disorder | 1 |
| Hereditary hemorrhagic telangiectasia | Vascular malformation disorder | 374 |
| Cerebral Cavernous Malformation | Vascular malformation disorder | 32 |
| Arteriovenous malformation | Vascular malformation disorder | 13 |
| Capillary malformation-arteriovenous malformation | Vascular malformation disorder | 9 |
| Lymphedema | Vascular malformation disorder | 8 |
| Klippel Trenaunay syndrome | Vascular malformation disorder | 5 |
| Lymphatic malformation | Vascular malformation disorder | 4 |
| Vascular malformation | Vascular malformation disorder | 3 |
| Venous malformation | Vascular malformation disorder | 2 |
| Blue rubber bleb nevus syndrome | Vascular malformation disorder | 1 |
| Carotid-Cavernous Fistula | Vascular malformation disorder | 1 |
| Maffucci syndrome | Vascular malformation disorder | 1 |
| Venous insufficiency | Vascular malformation disorder | 1 |

